# Health economic evaluation of lung cancer screening using a diagnostic blood test: the Early detection of Cancer of the Lung Scotland (ECLS)

**DOI:** 10.1101/2024.04.19.24306080

**Authors:** Jose Antonio Robles-Zurita, Nicola McMeekin, Frank Sullivan, Frances S Mair, Andrew Briggs

## Abstract

**Background:** Diagnostic blood tests have the potential to identify lung cancer in people at high risk, which is important as detecting lung cancer at an early stage is associated with survival advantages. We conducted an economic evaluation to assess the cost-effectiveness of a lung cancer screening intervention, using the EarlyCDT®-Lung Test with subsequent x-ray and low-dose chest CT scans (LDCT) for patients with a positive test result, compared to both usual care and LDCT screening for the whole target population.

**Methods:** A lifetime analyses with a UK NHS and personal social services perspective was conducted using a decision model for a target population of 1,000 individuals, where model parameters came from the ECLS study and literature. The model simulated the probability distribution of stage at cancer detection (early vs. late) for each evaluated alternative. Quality adjusted life-years (QALY) assigned to patients were dependant on stage at detection during or after the screening period, costs were dependent on the diagnostic pathway followed by patients and on cancer stage at diagnosis. We estimated net monetary benefit (NMB) at policy relevant cost-effectiveness thresholds for base-case, deterministic sensitivity, and scenario analyses.

**Results:** The base case incremental NMB of the ECLS intervention compared to *no screening* was £33,179 (95% CI: -£81,396.4, £147,180) and £140,609 (95% CI: £36,255.1, £316,612) respectively for a cost-effectiveness threshold of £20,000 and £30,000 per QALY. The same figures compared with *LDCT screening* were £162,095 (95% CI: £52,698.3, £271,735) and £52,185 (95% CI: -£113,152, £220,711). A deterministic sensitivity analysis indicated parameter values that resulted in a change to cost-effectiveness results, for example: prevalence of lung cancer (1%-4%); relative prevalence of early stage lung cancer (25%-75%); cost of the EarlyCDT®-Lung Test (£59-£201.5); test sensitivity for early stage lung cancer (25%-75%); and specificity of the test (50%-100%). A scenario analysis confirmed that the EarlyCDT®-Lung Test performs better than a zero-cost random test and showed that if the sensitivity of the test is assumed 25% (rather than base case 52%) the ECLS intervention would be not cost effective at a £30,000 per QALY threshold.

**Limitations:** Lack of trial resource data for the within study analysis resulted in partial reliance on expert opinion. Some participants may have modified their smoking behaviour due to participation in the trial.

**Conclusions:** The base case analysis results estimated that the ECLS intervention is the most cost-effective screening alternative, with highest probability of being cost-effective, when compared to no screening or LDCT screening. This result may change with modifications of the parameters, *prevalence of lung cancer* and *EarlyCDT®-Lung Test cost*, suggesting that the three alternatives considered in the main analysis are potentially cost-effective depending on the disease risk of the target population and the cost of testing.

## 1 Introduction

Lung cancer (LC) has the highest mortality of cancers worldwide[1], with Scotland having one of the highest rates in Scotland.[2] Early detection and diagnosis improves prognosis, the 5-year survival rate is approximately 60% for stage I LC but only 3% for those with stage IV.[3] 85% of LC cases are not diagnosed until they become symptomatic, at which point the cancer is advanced.[2] Early detection not only improves prognosis but the costs of treating early stage LC is around half that of treating stage IV cancer, evidence shows that achieving earlier diagnosis would be highly cost-effective.[4] ‘In 2022 the UK National Screening Committee recommended that all four nations implement targeted screening for lung cancer.[5] Whilst The Targeted Lung Health Checks programme currently running in England is a good starting point, screening is not yet a national programme implementation is under way in some places and more work is needed to achieve this.[6]

Effective screening programmes identify asymptomatic people with cancer, achieving early diagnosis.[4] Previous research in England showed screening using low-dose chest CT (LDCT) results in 85% of LC cases detected at stage I or II, over 90% of these cases are potentially curable with treatment.[7] However, CT scans are expensive and have been found to result in a high number of false positives (over 90% of tumours detected are benign), overdiagnosis and exposure to radiation.[8] Another drawback of LDCT screening programs is capacity; there is presently not the infrastructure to carry out large numbers of LDCT in the UK, which is why the targeted NHS lung cancer screening programme in England is not projected to be universally available until 2029.[9, 10]

The EarlyCDT®-Lung Test is a blood test that identifies biomarkers useful for the prediction of LC. The test detects early and late stages of LC with sensitivity of 41% and specificity of 90%[11], and could act as the first step in a targeted approach to LC screening for early stage detection. The Early detection of Cancer of the Lung Scotland (ECLS) trial designed a screening strategy where the target population was administered with the EarlyCDT®-Lung Test and, if tested positive, followed by chest x-ray and serial chest LDCT scanning.

The ECLS trial sought to assess the effectiveness and cost-effectiveness of this screening strategy, evaluating whether using the test, followed by chest x-ray and serial chest LDCT scanning, reduces incidence of patients with late stage LC (III & IV) or unclassified presentation (U) at diagnosis, compared to standard clinical practice (i.e. no-screening for LC).[12] After the first published results, some questions have been raised regarding the interpretation of the findings and the study design. First, ECLS found a difference in prevalence of LC between arms, which may contradict the intuitive expectation that screening interventions should not affect underlying incidence of LC in the target population.[12] Even if the reported difference in prevalence was due to chance alone; ideally, we should evaluate the screening intervention assuming equal prevalence in both treatment and comparator. Secondly, it has been suggested that the ECLS study may have underestimated LC prevalence overall, and therefore overestimated the sensitivity of the EarlyCDT®-Lung test.[13] The same source suggested that a more appropriate comparator for the new screening intervention could have been an LDCT only screening (i.e. the same as the intervention screening without administering the EarlyCDT®-Lung Test) rather than using standard clinical practice (no screening) [13]. Third, resource use data collected for intervention costs are only available for test positive participants which makes it necessary to use assumptions for participants with a negative test result.

In this paper, we use a modelling approach, that allows us to address the aforementioned issues, to analyse the cost-effectiveness of the proposed screening strategy (EarlyCDT®-Lung Test and, if tested positive, followed by chest x-ray and serial chest LDCT scanning) compared to no-screening detection, on the one hand, and LDCT scanning for the whole target population, on the other hand.

## 2 Methods

### 2.1 The ECLS trial

Details of the ECLS study are reported in the protocol and main clinical results paper and described briefly here.[2, 12] Participants between 50 and 75 years and at high risk of LC were recruited between April 2013 and July 2016. High risk was defined as current or former smokers with a minimum of 20 pack-years, or less than 20 pack-years plus a family history (parent, sibling, or child) of LC. Participants were healthy enough to undergo radical treatment either by pulmonary resection or stereotactic radiotherapy. It was expected that about 2% of participants would develop LC in the 24-months follow-up period of the trial based on a previous screening study with a similar target population.[14]

Participants were recruited from targeted general practices serving patients in the lowest quintile of deprivation in Scotland (from Greater Glasgow and Clyde, Tayside and Lanarkshire), as measured by the Scottish Index of Multiple Deprivation (SIMD).[15] Additional recruitment was attained through adverts, posters, flyers and community-based interactions. Potential participants were also recruited at the local clinical research centre from Tayside, Greater Glasgow and Clyde and Lanarkshire health boards. 12,209 participants were randomised to either the intervention or the control group and followed-up for 24 months.

The intervention (screening) arm comprised an EarlyCDT®-Lung Test administered to all subjects in the target population. The test result could be negative, in which case no additional investigation was offered, or positive, in which case immediate investigation by x-ray and LDCT imaging was offered. Participants in the screening arm with a positive test result and no evidence of LC from the x-ray and LDCT scan were invited for subsequent 6-monthly LDCT scans over the 24-months follow-up. Alternatively, if the results of the x-ray or LDCT scan were suspicious, contrast enhanced staging CT was undertaken, depending on the results of this scan the participant was referred to NHS care (clinically significant results) or continued with 6-monthly scans. The comparator (no-screening) arm comprised UK standard clinical practice at that time; awaiting the development of symptoms and investigation of those symptoms according to national guidelines.[16, 17] For outcomes, validated data on cancer occurrence, mortality and comorbidities were obtained, with patient consent, from National Services Scotland, a high-quality health services data repository. These were deterministically linked to baseline and follow-up visit data in OpenClinica (a clinical research service provider) using Scotland’s Community Health Index number and analysed in the Dundee Health Informatics Centre Safe Haven.[18] Pathology and tumour staging reports were prepared by independent assessors, blinded to the allocation status of study participants. Staging data were taken from the Scottish Cancer Registry (SMR06).[19]

### 2.2 Overview of economic analysis

The analysis used a NHS and personal social services perspective following recommendations by the National Institute for Health and Care Excellence (NICE), including healthcare costs only.[20] A 3.5% discounting rate was applied to outcomes and costs occurring after the first year from start of screening, 0% and 6% were used in a sensitivity analysis. Best practice methods and Consolidated Heath Economic Evaluation Reporting Standards (CHEERS) reporting guidelines were followed as appropriate.[21–23] A cost-effectiveness analysis was performed to estimate the incremental costs, quality adjusted life years (QALYs) and net monetary benefit using a decision tree model.

The model used information obtained by the ECLS trial about the LC stage at detection to extrapolate long-term QALYs and healthcare costs of LC patients. The key hypothesis behind the model is that a screening intervention which detects LC at an earlier stage will bring future benefits in terms of higher life expectancy (because the patient could benefit from earlier treatment), and lower treatment costs, whilst late-stage cancers are related to lower life expectancy and more resource use and costly treatments.

A decision analytic modelling approach is appropriate in the context of this study not only to estimate long term benefits but to overcome some challenges raised by the ECLS trial results. For example, the model allowed us to equalise LC prevalence in the screening intervention and the comparator as it is expected that screening interventions do not affect underlying incidence of LC in the target population.[12] Also, model parameters like the prevalence of LC and accuracy of the test could be modified to study plausible scenarios and alternative screening interventions, e.g. LDCT screening offered to all members of the target population.[13]

### 2.3 The model

The model shown in Figure 1 simulated the diagnostic pathway (type of screening tests or investigations administered to the individual), and LC stage at detection (LC status and stage at detection), for a cohort of participants in the ECLS screening intervention. Lifetime costs and QALYs were assigned to participants depending on the diagnosis pathway and disease stage at detection.

**Fig 1.**
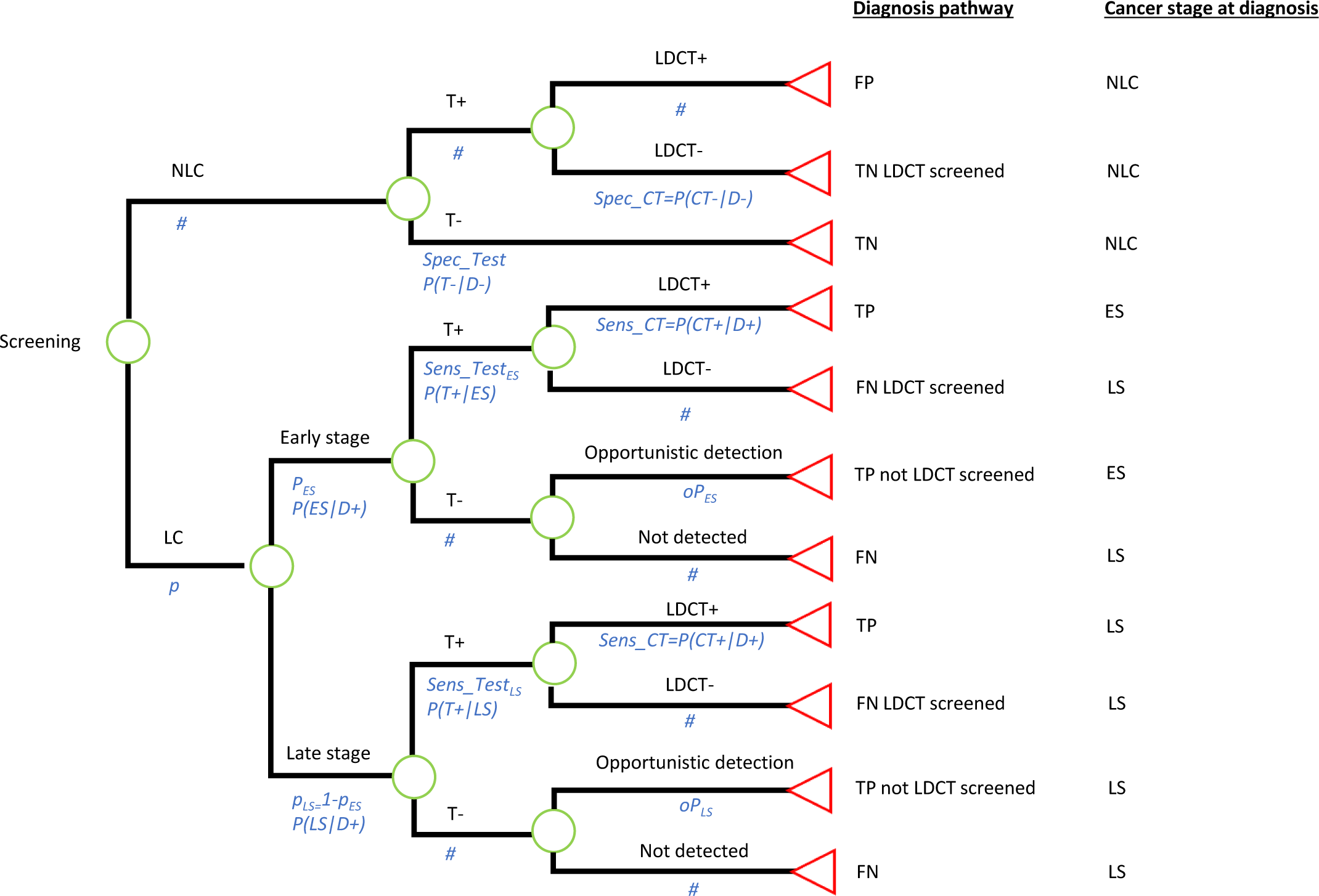
Decision tree model to simulate screening intervention and comparators: a) ECSL trial intervention, if specificity and sensitivity are those of the EarlyCDT®-Lung Test; b) Comparator 1: Standard care (no screening), if specificity is set at 1 and sensitivity is set at 0; c) Comparator 2: LDCT screening to all individuals in the target population, if specificity is set at 0 and sensitivity is set at 1. TN – true negative; TP – true positive; FN – false negative; FP – false positive; LDCT – low-dose computed tomography; LC – lung cancer; NLC – no lung cancer; ES – early stage; LS – late-stage.

#### 2.3.1 Structure and endpoints: disease pathway and disease stage at detection

An underlying LC prevalence (𝑝) of LC at the start of the screening intervention was assumed to be either early stage (I/II) or late stage (III, IV or U) with conditional probabilities 𝑝_𝐸𝑆_and 𝑝_𝐿𝑆_respectively. Participants undergo the EarlyCDT®-Lung test with specificity given by the probability of correctly identify disease-negative participants, i.e. P(T^-^|D^-^). A different sensitivity can be specified for early stage (ES) and late stage (LS) cancers given by the probabilities P(T^+^|ES) and P(T^+^|LS). Participants with a positive test result are sent to 6-monthly LDCTs as designed in the ECLS trial, and those with a negative test result are no longer investigated. LDCT investigations could confirm a LC diagnosis or not depending on the true disease state of the patient and the sensitivity and specificity of the 6-monthly LDCTs. Finally, LC patients who did not undertake LDCT investigations could be detected opportunistically during the screening period, for example in a hospital visit related or unrelated to LC symptoms, with probabilities 𝑂𝑝_𝐸𝑆_and 𝑂𝑝_𝐿𝑆_for ES and LS respectively.

The model classified individuals depending on their diagnosis pathway:

- True positive (TP). Individuals with LC that obtained a positive test result and were correctly identified by LDCTs.

- TP (not LDCT screened). Individuals with LC that obtained a negative test result and therefore were not offered LDCTs. These cases were opportunistically detected.

- True Negative (TN). Individuals with no LC that obtained a negative test result.

- TN (LDCT screened). Individuals with no LC that obtained a positive test result and were investigated by LDCTs with negative results.

- False Positive (FP). Those individuals with no LC but with a positive result from the test and from LDCT investigations.

- False Negative (FN). Those individuals with LC that obtained a negative test result.

- FN (LDCT screened). Those individuals with LC that obtained a positive test result but received a negative result after LDCT investigations.

The model classified individuals according to LC stage at diagnosis, into:

H^No LC. Individuals that were disease negative during the period of the ECLS screening intervention.

- ES LC. Individuals that had LC at ES during the screening intervention and were correctly detected during the course of the screening intervention by LDCTs. These patients were assumed to be detected at early stage and therefore to be benefited by early treatment. They were attached a high(er) life expectancy, consistent with being detected soon(er).

- LS LC. Two subgroups of patients can be considered. First, individuals that were at LS in the screening period. Second, individuals that had ES LC during the screening period but were undetected, either because they had a negative test result with no opportunistic detection, or because they had a positive test result followed by a LDCT negative result. The LS LC patients were assumed to be diagnosed too late to benefit from early treatment. Hence, they were attached a low(er) life expectancy.

In the analysis, all the disease-pathway groups were assigned the same cost for the administration of the test. However, the cost of LDCTs and other diagnostic imaging were different depending on the specific pathway followed by each group of individuals. For example, a *TN (LDCT screened)* participant incurred LDCT screening costs while a *TN* individual was not investigated due to a negative test result. Also, treatment costs were dependent on the LC stage at detection to account for the differences in type of therapies administered. Finally, life time and health utilities, used to construct QALYs, were conditional on the cancer stage at moment of diagnosis to account for the fact that quality of life and life expectancy is lower for LS cases.[3]

#### 2.3.2 ECLS screening intervention and comparators

The model depicted in Figure 1 allowed us to simulate the alternatives compared in this analysis by changing the sensitivity and specificity of the test provided before the administration of LDCT screening:

- <u>Intervention:</u> The ECLS intervention. This strategy was simulated by using the sensitivity and specificity of the EarlyCDT®-Lung Test as estimated in the ECLS trial published results.[12] Under this alternative, only some patients will obtain a positive test result and therefore only some of them will be screened by 6-monthly LDCTs for 2 years.

- <u>Comparator 1:</u> No screening. This strategy was simulated by assuming a test with sensitivity 0 and specificity 1. Therefore, none of the participants will obtain a positive test result and none of them will be sent to LDCT screening. The test used in this strategy would have a zero cost.

- <u>Comparator 2:</u> LDCT screening administered 6-monthly for 2 years. This strategy was simulated by setting the sensitivity and specificity of the test to 1 and 0, respectively. In this case, all the subjects in the target population will be sent to LDCT screening. Again, this test would be administered at no cost.

#### 2.3.3 Parameters: sources and estimation

The model parameters are listed in Table 1, including pathway probabilities, costs and outcomes for base case, probabilistic and deterministic sensitivity analysis (PSA and DSA). Details of parameters estimation are found in the *supplementary material*.

**Table 1:**
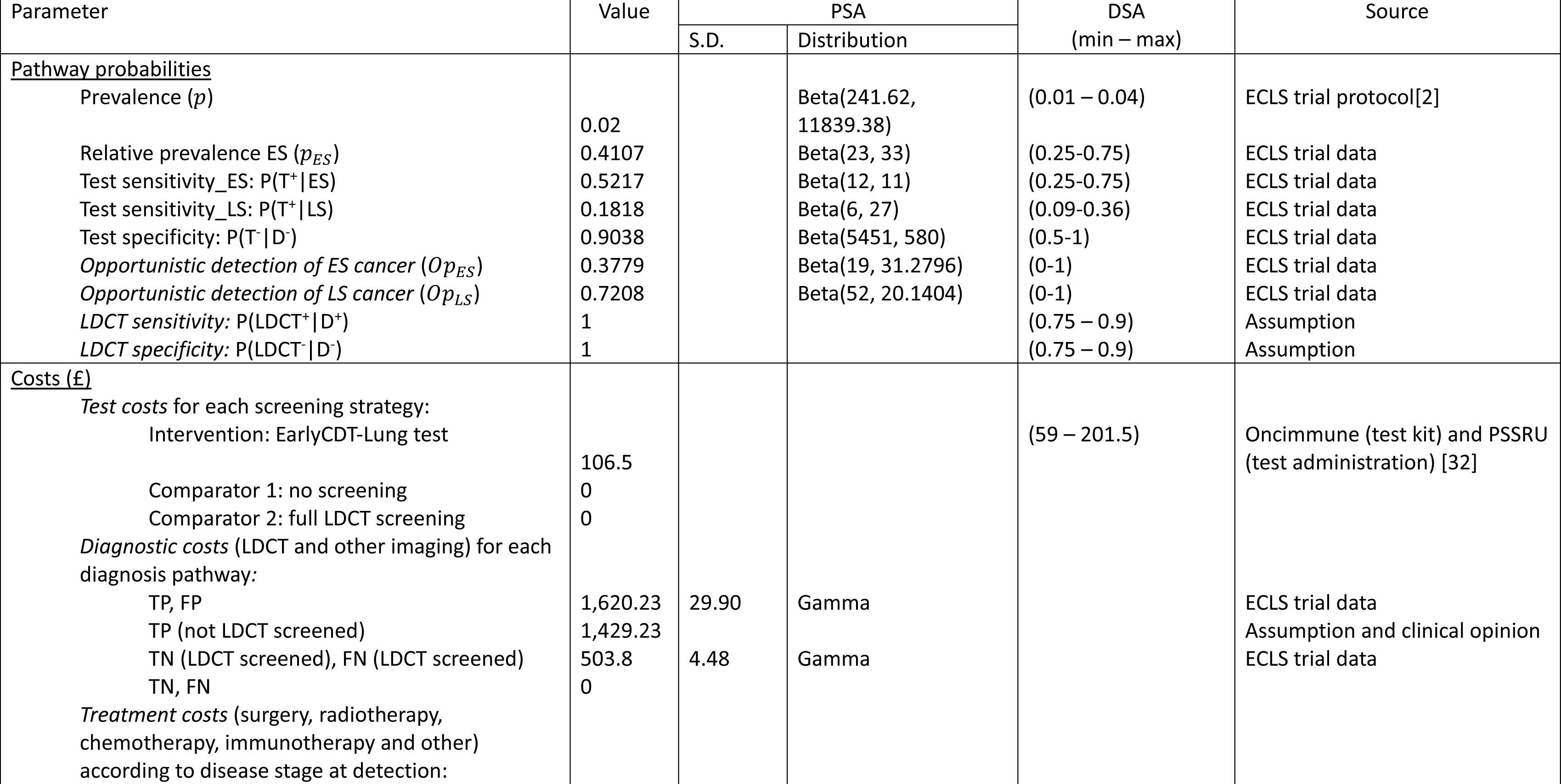
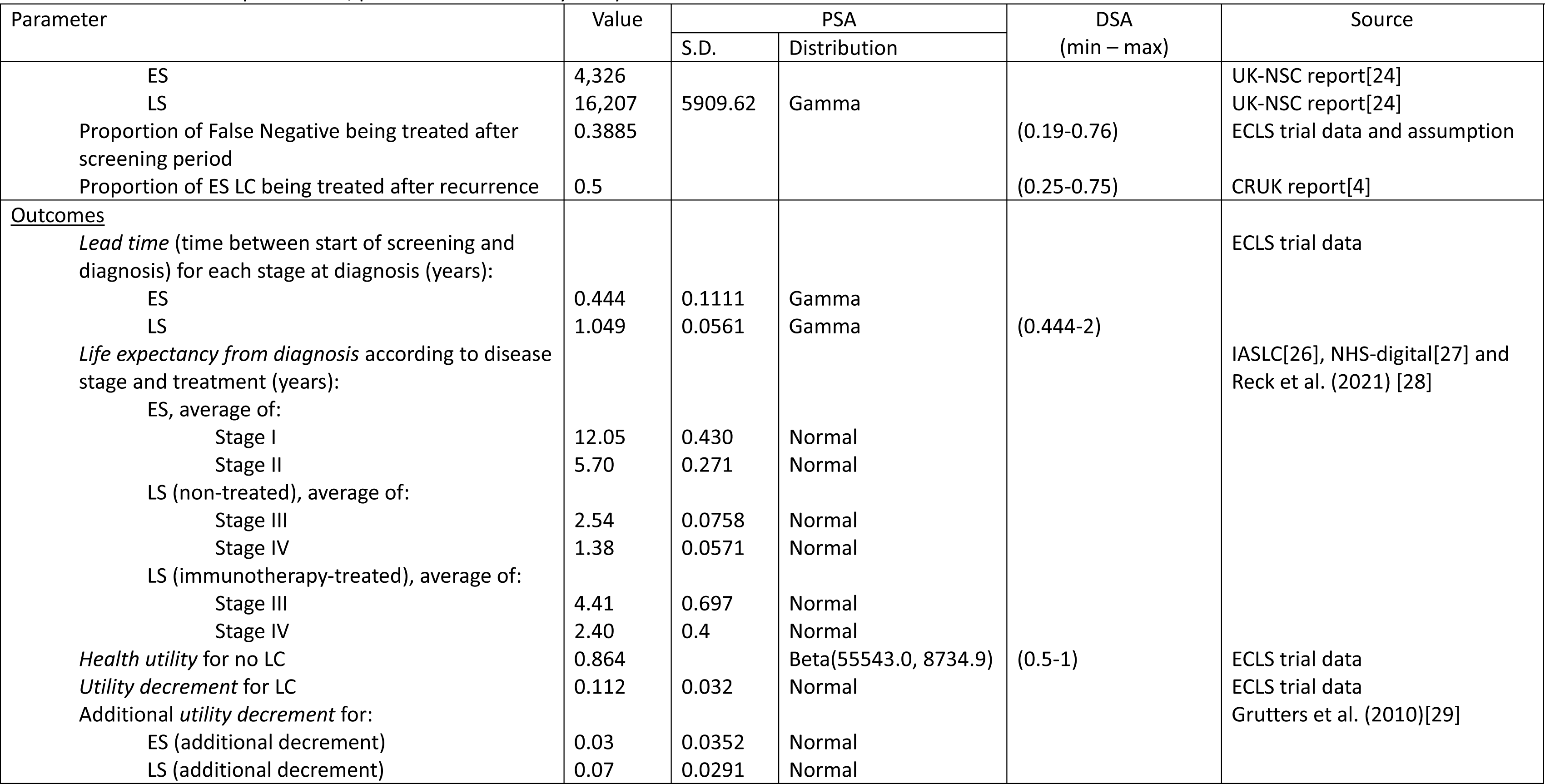
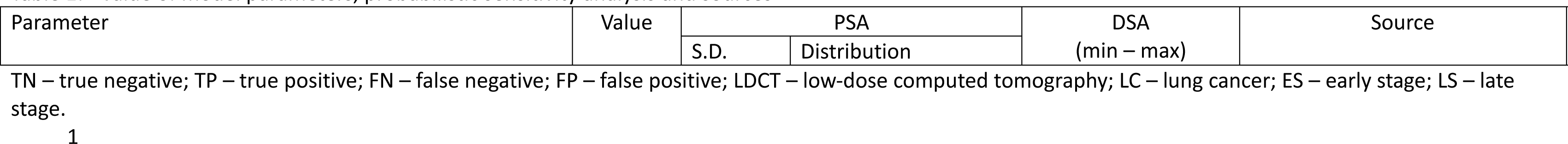
– Value of model parameters, probabilistic sensitivity analysis and sources

The base case analysis used a prevalence (*p*) of LC of 2% following the study protocol expectations based on a previous study of a similar population.[14] In a sensitivity analysis, prevalence was changed to 1% and 4% to relax this assumption. All the remaining probabilities were estimated using the data collected in the ECLS trial. The relative prevalence of ES cancers (𝑝_𝐸𝑆_) was estimated as the ratio of expected total ES cases to LS cases in the ECLS trial intervention arm using data of the cases detected and sensitivity of the EarlyCDT®-Lung Test for each LC stage. Sensitivity of the EarlyCDT®-Lung Test, for ES and LS cancers, and specificity was estimated as reported in the ECLS trial published manuscript.[12] The probability of opportunistic detection (𝑂𝑝_𝐸𝑆_and 𝑂𝑝_𝐿𝑆_) was estimated using data on detected LC in the ECLS trial control (no-screening) arm. Finally, the 6-monthly LDCT screening for two years was assumed to have a sensitivity and specificity of 100% in the base case analysis, i.e. no false positive or false negative were allowed for participants sent to LDCT screening.

Costs parameters represented 2021/22 pounds sterling and comprised the administration of a LC test, i.e. the EarlyCDT®-Lung Test for the intervention alternative, diagnostic costs for LDCT screening and imaging, and treatment costs. Costs of the test were applied at the beginning of the screening programme. However, diagnostic costs were assumed to occur one year after (i.e. the average time for a two-year screening programme) or at moment of diagnosis if the person was diagnosed during the course of the screening intervention.

Treatment costs were assumed to happen in the same year of diagnosis. The cost of the EarlyCDT®-Lung Test kit was advised by Oncimmune (£95) and its administration was assumed equivalent to 15 minutes of nurse time at GP surgery (£11.50). The testing costs were applied only to individuals in the ECLS intervention. The diagnostic costs differed by diagnostic pathway. Average number of x-ray and CT scans were estimated from ECLS data and valued for patients with a positive test result (i.e. those sent to LDCT screening) differentiating between those with and without a confirmatory LC diagnosis. In addition, a confirmatory diagnostic cost was applied to all participants with a diagnosis of LC. Confirmatory diagnostic tests were based on opinion of clinical experts in the ECLS study and consisted of an x-ray, a contrast CT scan and either a bronchoscopy or CT guided biopsy with the average of a bronchoscopy and a CT guided biopsy cost applies. Individuals with a negative test result and no diagnosis had a zero diagnostic cost. Lung cancer treatment costs included surgery, radiotherapy, chemotherapy, immunotherapy and other as calculated in a UK National Screening Committee (UK-NSC) report authored by the Exeter Test Group and Health Economics Group.[24] Average treatment costs at diagnosis for LC stage I and II, as calculated by the UK-NSC report, was assigned to ES. In the same way, average treatment costs at diagnosis for LC stage III and IV was assigned to LS. Only 38.85% of LS LC not detected during the screening period will be expected to receive treatment before dying according to the opportunistic detection rate estimated with the ECLS study data. Also, a proportion of ES LC were assumed to recur and incur in lung cancer treatment for advanced stage (about 50% according to Cancer Research UK).[4] Unit costs and details of computation of each cost item are in the *supplementary material*.

The computation of QALYs for participants in the screening programme involved the multiplication of lifetime (LT) and health utility (HE). LT for LC patients was computed as the addition of two constructs: 1) *lead time*, defined as the time passed between start of the screening programme and diagnosis, and 2) life expectancy (LE) from diagnosis. Lead time for TP LC detected at ES was estimated as the average for LC cases with a positive test result in the ECLS trial intervention arm, with a mean of 0.444 years. For consistency, the same value was attached to TP (not LDCT screened) early-stage LCs. Late-stage LCs were assigned the average lead time for LC cases in the control arm and LC cases with a negative test result in the intervention arm, which was 1.049 years. These lead time estimations imply an average time to progression (from early to late stage) of about 0.6 years, which lies within the range of previous estimations for Caucasian patients.[25] Weibull survival curves were estimated to compute life expectancy from diagnosis using digitized data from K-M curves published by the International Association for the Study of LC IASLC staging project. [26] Then survival parameters were calibrated for each LC stage using five-year survival figures from an England national study that followed lung cancer patients diagnosed from 2015 to 2019 and followed up to 2020.[27] Finally, the model assigned the average LE of LC stage I and II to ES patients. LE of stage III and IV, calibrated from English data, is unlikely to capture the effect of immunotherapy on survival of advanced lung cancer, recommended from the year 2019, but rather it is the result of previous standard treatments such as chemotherapy. A modified LE was computed for LS LC receiving immunotherapy by applying a hazard ratio estimated by the KEYNOTE-24 study (HR=0.62, 95% CI: 0.48 to 0.81; overall survival of immunotherapy vs. chemotherapy).[28] Immunotherapy-augmented life expectancy was applied to LS LC detected during or after the screening period. On the contrary, “pre-immunotherapy” survival figures were assigned to those patients expected to die before receiving treatment. An average health utility for patients with and without LC was estimated from EQ-5D responses reported in the ECLS study at baseline. Previous literature was followed by attaching a decremental health utility to each stage.[29] Details of the estimation and computation of lead time, life expectancy and health utilities are found in the *supplementary material*.

### 2.4 Sensitivity and scenario analysis

A PSA was conducted via 10,000 Monte Carlo simulations using the following distribution functions: -beta distribution for prevalence, sensitivity, specificity, probability of opportunistic detection and health utilities; - normal distribution for life expectancy and incremental health utilities; - gamma distribution for lead time bias, diagnostic costs for the test positive patients, and late-stage treatment costs. Remaining parameters based on deterministic data, assumptions, or expert opinion, were fixed: treatment costs for ES LC; cost of EarlyCDT®-Lung Test; and diagnostic costs for test negative patients. Also, a DSA was conducted, to check the robustness of the results to changes in key parameters of the ECLS intervention (among them: prevalence of lung cancer in the target population, relative prevalence of ES LC, cost of the test, sensitivity, and specificity) and to illustrate the mechanism of the model. The minimum and maximum values used for the DSA for key parameters were chosen to make an impact on the cost-effectiveness results at the policy relevant thresholds.

Two scenario analyses were conducted to address some challenges relating to the interpretation of the ECLS study. First, sensitivity of EarlyCDT®-Lung Test was set to 25%, addressing concerns about overestimation of sensitivity of the test for early LCs.[13] Second, the false positive rate (1-specificity) was set equal to the true positive rate (sensitivity) and cost of the test to zero. The latter simulates a strategy where patients receive a completely random test; patients are sent to LDCT screening independently of their true disease state with a probability of 52%.

### 2.5 Cost-effectiveness analysis

Mean and 95% normal confidence intervals for the PSA were estimated for costs and QALYs for the intervention and the two comparators for each 1,000 participants. Net Monetary Benefit (NMB) was also estimated using policy relevant cost-effectiveness thresholds (20,000 and 30,000 GBP per QALY)[20] and cost effectiveness acceptability curves (CEACs) calculated. Results of the DSA were presented as mean costs, QALYs and NMBs. The scenario analysis was presented on a cost-effectiveness plane for ease of interpretation.

The statistical analysis has been performed using STATA 18.0 (StataCorp, TX, USA) and the model has been run in R using the package *heemod*.[30, 31]

## 3 Results

### 3.1 Base case analysis

Table 2 includes QALYs, costs and NMB per 1,000 participants for the base case analysis. NMB figures point to the ECLS intervention as the most cost-effective alternative. The incremental NMB estimates of the ECLS intervention compared to *no screening* are subject to some uncertainty with £33,179 (95% CI: -£81,396.4, £147,180) and £140,609 (95% CI: £36,255.1, £316,612) for a cost-effectiveness threshold of £20,000 and £30,000 per QALY respectively. The ECLS intervention brings both higher costs, £181,681 (95% CI: £168,243, £195,121), and QALYs, 10.7 (95% CI: 4.5, 17), at a ratio of less than £20,000 per QALY. The use of the ECLS intervention implies 2.67 more early-stage LCs than in the no screening alternative at the expense of 94.2 participants (out of 980 of no LC individuals) being unnecessarily investigated (see *supplementary material* for model counts for each diagnostic pathway).

**Table 2.** Base case cost-effectiveness results: costs, QALYs and NMBs per one thousand participants.

|  | ECLS intervention |  | Comparator 1: No screening |  | Comparator 2: LDCT screening |  |
| --- | --- | --- | --- | --- | --- | --- |
|  | Mean | [95% CI] | Mean | [95% CI] | Mean | [95% CI] |
| Costs (£) | 414,102 | [376,719, 451,792] | 232,421 | [196,659, 268,489] | 796,016 | [755,178, 837,512] |
| QALYs | 8,570.4 | [8,086.9, 9,050] | 8,559.7 | [8,076.8, 9,038.7] | 8,581.4 | [8,097.3, 9,061.6] |
| NMB ($\lambda$=£20,000) | 170,994,000 | [161,323,000, 180,586,000] | 170,961,000 | [161,302,000, 180,541,000] | 170,832,000 | [161,148,000, 180,437,000] |
| NMB ($\lambda$=£30,000) | 256,698,000 | [242,192,000, 271,085,000] | 256,557,000 | [242,069,000, 270,928,000] | 256,645,000 | [242,121,000, 271,052,000] |
| ΔCosts (£) | | | 181,681 | [168243, 195121] | -381,915 | [-401,080, -363,100] |
| ΔQALYs | | | 10.7 | [4.5, 17] | -11.0 | [-16.8, -5.16] |
| ΔNMB ($\lambda$=£20,000) | | | 33,179 | [-81396.2, 147180] | 162,095 | [52,698.4, 271,735] |
| ΔNMB ($\lambda$=£30,000) | | | 140,609 | [-36255, 316612] | 52,185 | [-115,152, 219,711] |

CI – Confidence interval; $\lambda$ – Cost effectiveness threshold; QALYs – Quality adjusted life years; NMB – Net monetary benefit; monetary amounts rounded to six significant figures and no decimal places; QALYs rounded to one decimal place

The incremental NMB compared to *LDCT screening* is £162,095 (95% CI: £52,698.3, £271,735) and £52,185 (95% CI: -£113,152, £219,711) respectively for the lower and higher threshold. Even though a LDCT screening would bring a gain of 11 QALYs (95% CI: 5.16, 16.831), due to more ES cancers detected, it would also incur £381,915 in higher costs (95% CI: £401,080, £363,100) because all no-LC participants would be sent for LDCT investigation. Out of the 20 patients (2% of 1,000) that would have LC in the analysis cohort, an average of 8.21 would be detected at an ES if investigated by LDCT, 2.44 more than in the case of the ECLS intervention. At the same time, a full LDCT screening would send 980 participants with no LC to unnecessary CT investigations, whereas only 94.2 would be tested positive with the EarlyCDT-Lung test. LDCT screening, compared to the ECLS intervention, would gain one QALY at a cost of £34,810.

The CEACs in figure 2 shows that the ECLS intervention has the highest probability of being cost-effective for any cost-effectiveness threshold between £18,000 and £35,000. Below £18,000 per QALY *no screening* would become the alternative with a higher probability to be cost effective. A LDCT screening would become the one with a higher probability of cost-effectiveness if we set a value of QALY above £35,000. The PSA concludes that the ECLS intervention has a maximum 82.39% probability of being cost-effective at a threshold of £25,000 per QALY.

### 3.2 Deterministic and scenario analyses

The deterministic sensitivity analysis is shown in Table 3 where we see how cost-effectiveness changes with the cost of the EarlyCDT test and with prevalence of LC in the target population. Reducing the cost of the test to half (£47.5 rather than £90, plus £11.50 administration costs) would further improve the cost-effectiveness of the ECLS intervention. However, if we double the cost of the EarlyCDT test (£190 plus £11.5 administration costs) LDCT screening would stop being the most cost-effective alternative; although the 95% CIs shows much uncertainty in this case. If prevalence was set to 1% *no screening* would be the most cost-effective alternative, even though the ECLS intervention would continue being better than a LDCT screening. If prevalence was as high as 4%, LDCT screening would be the most cost-effective alternative at the two thresholds used. The proportion of early-stage cases among all the lung cancers is also a relevant factor; for example, if this proportion was 25% then the ECLS screening would not be cost-effective at a threshold of £20,000 per QALY, favouring *no screening*. On the other hand, a 75% relative prevalence of ES LC would favour *full LDCT screening* as the best strategy at a threshold of £30,000 per QALY. The ECLS intervention would not be cost-effective if the test sensitivity for ES LC was only 0.25, or if the specificity was as low as 0.5, at any policy relevant thresholds. The DSA (in supplementary document) shows that cost-effectiveness of the three alternatives compared could be affected by: the rate of opportunistic detection for ES lung cancer (higher rate favouring *no screening* or ECLS intervention vs. full LCDT screening), discount rate (lower rate favouring the ECLS and LDCT screening interventions), health utilities used (lower base utilities favouring the *no screening* alternative), and LCDT sensitivity (lower sensitivity favouring *no screening*).

**Table 3.**
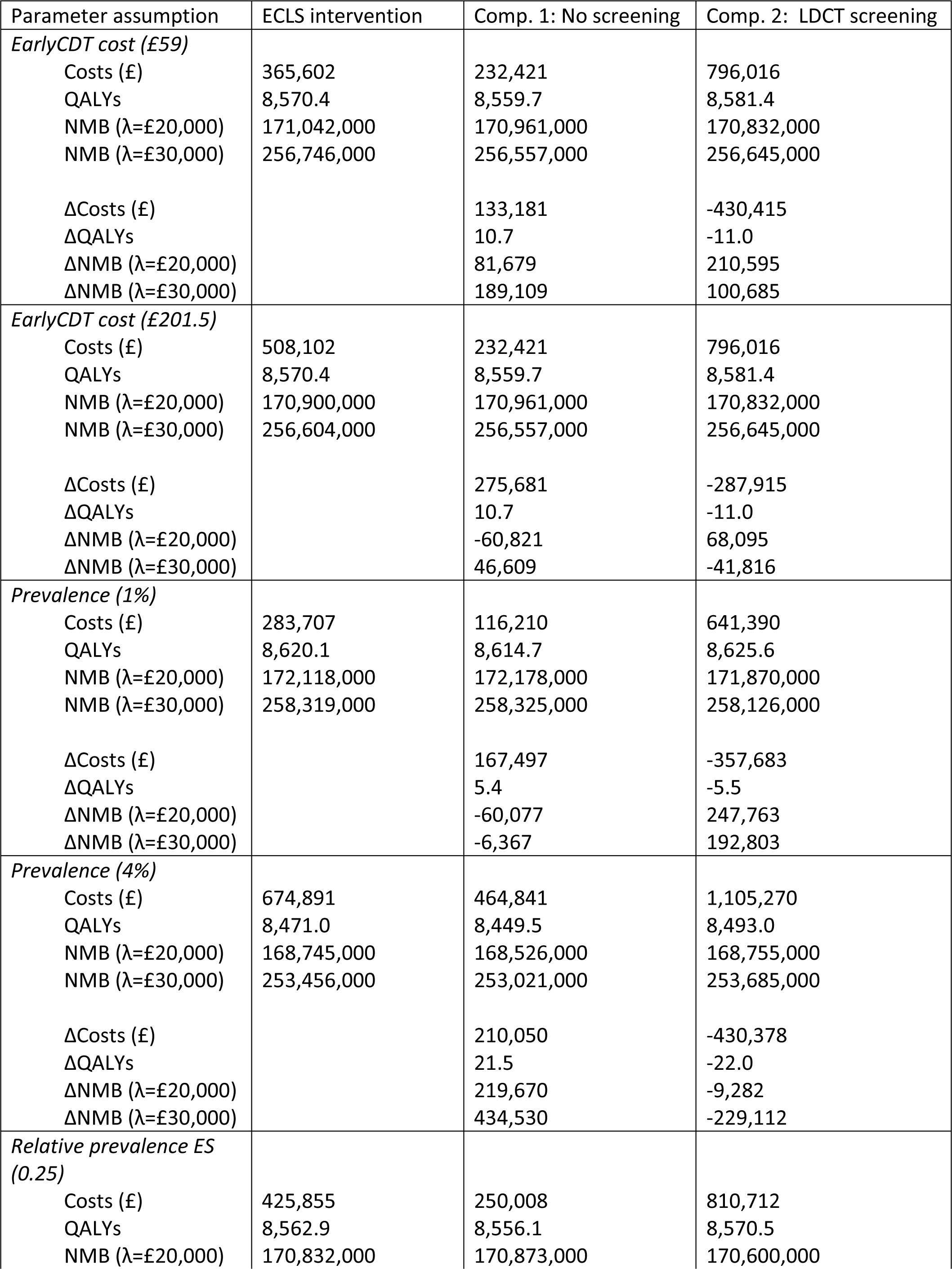
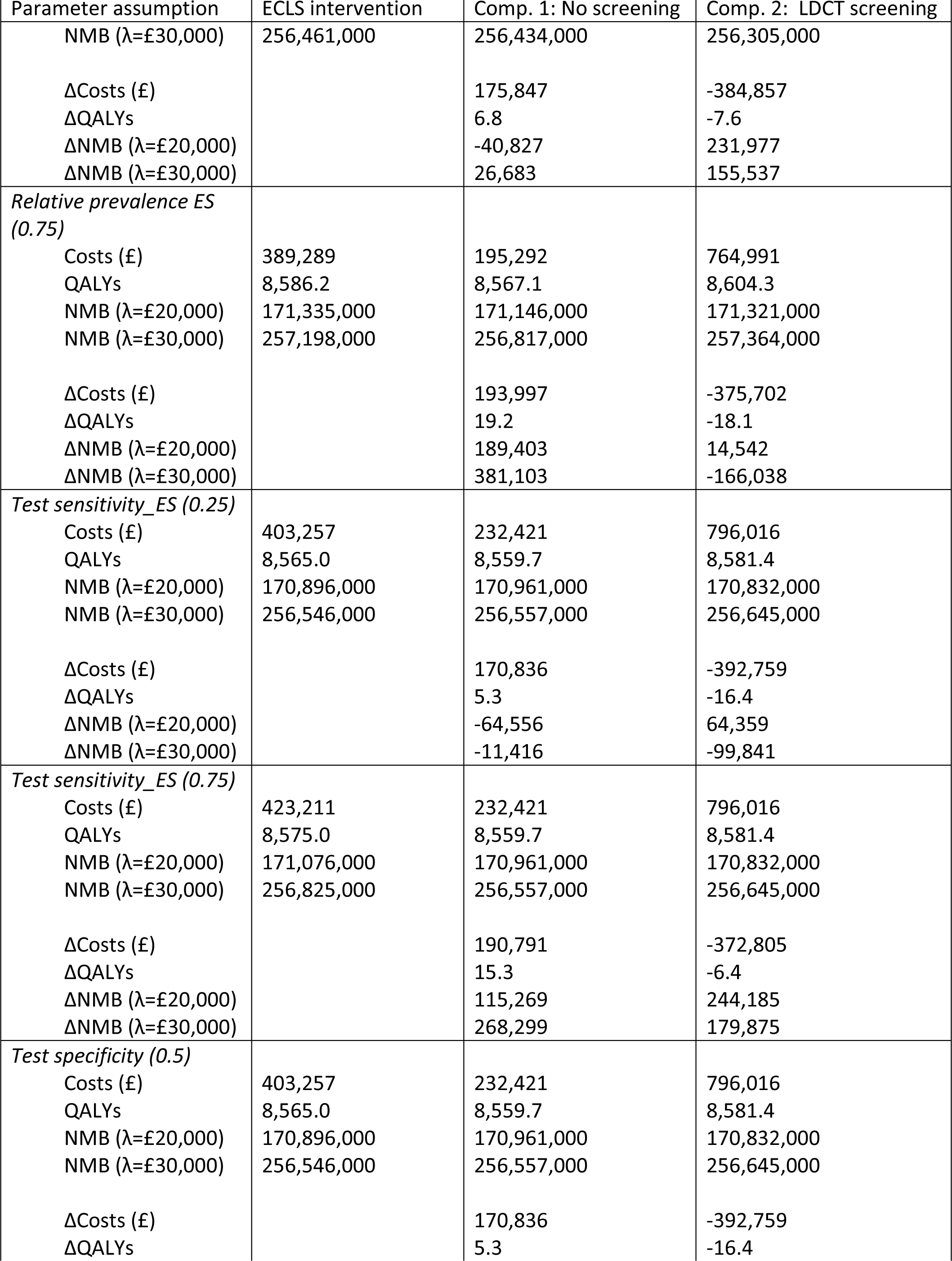
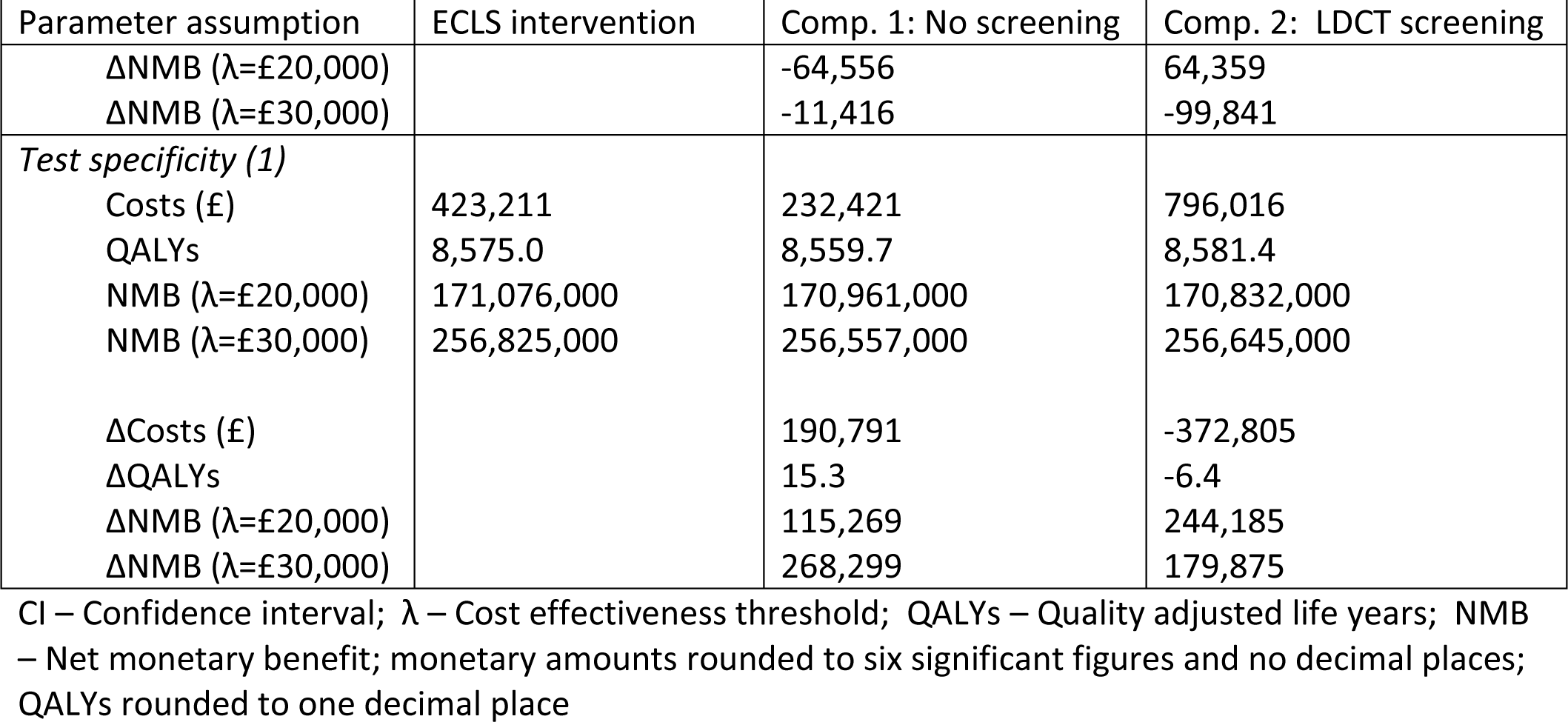
Deterministic sensitivity analysis: costs, QALYs and NMBs per one thousand participants.

| Parameter assumption | ECLS intervention | Comp. 1: No screening | Comp. 2: LDCT screening |
| --- | --- | --- | --- |
| *EarlyCDT cost (£59)* |  |  |  |
| Costs (£) | 365,602 | 232,421 | 796,016 |
| QALYs | 8,570.4 | 8,559.7 | 8,581.4 |
| NMB ($\lambda$=£20,000) | 171,042,000 | 170,961,000 | 170,832,000 |
| NMB ($\lambda$=£30,000) | 256,746,000 | 256,557,000 | 256,645,000 |
| ΔCosts (£) | | 133,181 | -430,415 |
| ΔQALYs | | 10.7 | -11.0 |
| ΔNMB ($\lambda$=£20,000) | | 81,679 | 210,595 |
| ΔNMB ($\lambda$=£30,000) | | 189,109 | 100,685 |
| *EarlyCDT cost (£201.5)* |  |  |  |
| Costs (£) | 508,102 | 232,421 | 796,016 |
| QALYs | 8,570.4 | 8,559.7 | 8,581.4 |
| NMB ($\lambda$=£20,000) | 170,900,000 | 170,961,000 | 170,832,000 |
| NMB ($\lambda$=£30,000) | 256,604,000 | 256,557,000 | 256,645,000 |
| ΔCosts (£) | | 275,681 | -287,915 |
| ΔQALYs | | 10.7 | -11.0 |
| ΔNMB ($\lambda$=£20,000) | | -60,821 | 68,095 |
| ΔNMB ($\lambda$=£30,000) | | 46,609 | -41,816 |
| *Prevalence (1%)* |  |  |  |
| Costs (£) | 283,707 | 116,210 | 641,390 |
| QALYs | 8,620.1 | 8,614.7 | 8,625.6 |
| NMB ($\lambda$=£20,000) | 172,118,000 | 172,178,000 | 171,870,000 |
| NMB ($\lambda$=£30,000) | 258,319,000 | 258,325,000 | 258,126,000 |
| ΔCosts (£) | | 167,497 | -357,683 |
| ΔQALYs | | 5.4 | -5.5 |
| ΔNMB ($\lambda$=£20,000) | | -60,077 | 247,763 |
| ΔNMB ($\lambda$=£30,000) | | -6,367 | 192,803 |
| *Prevalence (4%)* |  |  |  |
| Costs (£) | 674,891 | 464,841 | 1,105,270 |
| QALYs | 8,471.0 | 8,449.5 | 8,493.0 |
| NMB ($\lambda$=£20,000) | 168,745,000 | 168,526,000 | 168,755,000 |
| NMB ($\lambda$=£30,000) | 253,456,000 | 253,021,000 | 253,685,000 |
| ΔCosts (£) | | 210,050 | -430,378 |
| ΔQALYs | | 21.5 | -22.0 |
| ΔNMB ($\lambda$=£20,000) | | 219,670 | -9,282 |
| ΔNMB ($\lambda$=£30,000) | | 434,530 | -229,112 |
| *Relative prevalence ES (0.25)* |  |  |  |
| Costs (£) | 425,855 | 250,008 | 810,712 |
| QALYs | 8,562.9 | 8,556.1 | 8,570.5 |
| NMB ($\lambda$=£20,000) | 170,832,000 | 170,873,000 | 170,600,000 |

| Parameter assumption | ECLS intervention | Comp. 1: No screening | Comp. 2: LDCT screening |
| --- | --- | --- | --- |
| NMB ($\lambda$=£30,000) | 256,461,000 | 256,434,000 | 256,305,000 |
| ΔCosts (£) | | 175,847 | -384,857 |
| ΔQALYs | | 6.8 | -7.6 |
| ΔNMB ($\lambda$=£20,000) | | -40,827 | 231,977 |
| ΔNMB ($\lambda$=£30,000) | | 26,683 | 155,537 |
| *Relative prevalence ES (0.75)* |  |  |  |
| Costs (£) | 389,289 | 195,292 | 764,991 |
| QALYs | 8,586.2 | 8,567.1 | 8,604.3 |
| NMB ($\lambda$=£20,000) | 171,335,000 | 171,146,000 | 171,321,000 |
| NMB ($\lambda$=£30,000) | 257,198,000 | 256,817,000 | 257,364,000 |
| ΔCosts (£) | | 193,997 | -375,702 |
| ΔQALYs | | 19.2 | -18.1 |
| ΔNMB ($\lambda$=£20,000) | | 189,403 | 14,542 |
| ΔNMB ($\lambda$=£30,000) | | 381,103 | -166,038 |
| *Test sensitivity_ ES (0.25)* |  |  |  |
| Costs (£) | 403,257 | 232,421 | 796,016 |
| QALYs | 8,565.0 | 8,559.7 | 8,581.4 |
| NMB ($\lambda$=£20,000) | 170,896,000 | 170,961,000 | 170,832,000 |
| NMB ($\lambda$=£30,000) | 256,546,000 | 256,557,000 | 256,645,000 |
| ΔCosts (£) | | 170,836 | -392,759 |
| ΔQALYs | | 5.3 | -16.4 |
| ΔNMB ($\lambda$=£20,000) | | -64,556 | 64,359 |
| ΔNMB ($\lambda$=£30,000) | | -11,416 | -99,841 |
| *Test sensitivity_ ES (0.75)* |  |  |  |
| Costs (£) | 423,211 | 232,421 | 796,016 |
| QALYs | 8,575.0 | 8,559.7 | 8,581.4 |
| NMB ($\lambda$=£20,000) | 171,076,000 | 170,961,000 | 170,832,000 |
| NMB ($\lambda$=£30,000) | 256,825,000 | 256,557,000 | 256,645,000 |
| ΔCosts (£) | | 190,791 | -372,805 |
| ΔQALYs | | 15.3 | -6.4 |
| ΔNMB ($\lambda$=£20,000) | | 115,269 | 244,185 |
| ΔNMB ($\lambda$=£30,000) | | 268,299 | 179,875 |
| *Test specificity (0.5)* |  |  |  |
| Costs (£) | 403,257 | 232,421 | 796,016 |
| QALYs | 8,565.0 | 8,559.7 | 8,581.4 |
| NMB ($\lambda$=£20,000) | 170,896,000 | 170,961,000 | 170,832,000 |
| NMB ($\lambda$=£30,000) | 256,546,000 | 256,557,000 | 256,645,000 |
| ΔCosts (£) | | 170,836 | -392,759 |
| ΔQALYs | | 5.3 | -16.4 |

| Parameter assumption | ECLS intervention | Comp. 1: No screening | Comp. 2: LDCT screening |
| --- | --- | --- | --- |
| $\Delta\text{NMB } (\lambda=\text{£}20,000)$ | | -64,556 | 64,359 |
| $\Delta\text{NMB } (\lambda=\text{£}30,000)$ | | -11,416 | -99,841 |
| *Test specificity (1)* |  |  |  |
| Costs (£) | 423,211 | 232,421 | 796,016 |
| QALYs | 8,575.0 | 8,559.7 | 8,581.4 |
| NMB ($\lambda=\text{£}20,000$) | 171,076,000 | 170,961,000 | 170,832,000 |
| NMB ($\lambda=\text{£}30,000$) | 256,825,000 | 256,557,000 | 256,645,000 |
| $\Delta\text{Costs } (\text{£})$ | | 190,791 | -372,805 |
| $\Delta\text{QALYs}$ | | 15.3 | -6.4 |
| $\Delta\text{NMB } (\lambda=\text{£}20,000)$ | | 115,269 | 244,185 |
| $\Delta\text{NMB } (\lambda=\text{£}30,000)$ | | 268,299 | 179,875 |

CI – Confidence interval; $\lambda$ – Cost effectiveness threshold; QALYs – Quality adjusted life years; NMB – Net monetary benefit; monetary amounts rounded to six significant figures and no decimal places; QALYs rounded to one decimal place

Base-case and scenario alternatives are represented on a cost-effectiveness plane as incremental QALYs and costs compared to *no screening* in Figure 2. Three lines representing three value thresholds (£10,000, £20,000 and £30,000 per QALY) are depicted for interpretation of the results. The ECLS intervention is the most cost-effective alternative, below the £20,000 threshold. The scenario with EarlyCDT®-Lung Test sensitivity set to 25% is the least cost effective with an incremental cost effectiveness ratio (ICER) above £30,000 per QALY. A random test, sending 52% of participants to LDCT screening independently on LC state, would be cost effective (compared to *no screening*) at the £30,000 threshold, even though it would be much less costly than a full LDCT screening (i.e. sending 100% of participants to LDCT screening). Notice that a zero-cost random test would necessarily be at the same cost-effectiveness threshold than a full LDCT screening strategy, only the scale of costs and QALYs would be modified.

**Fig 2.**
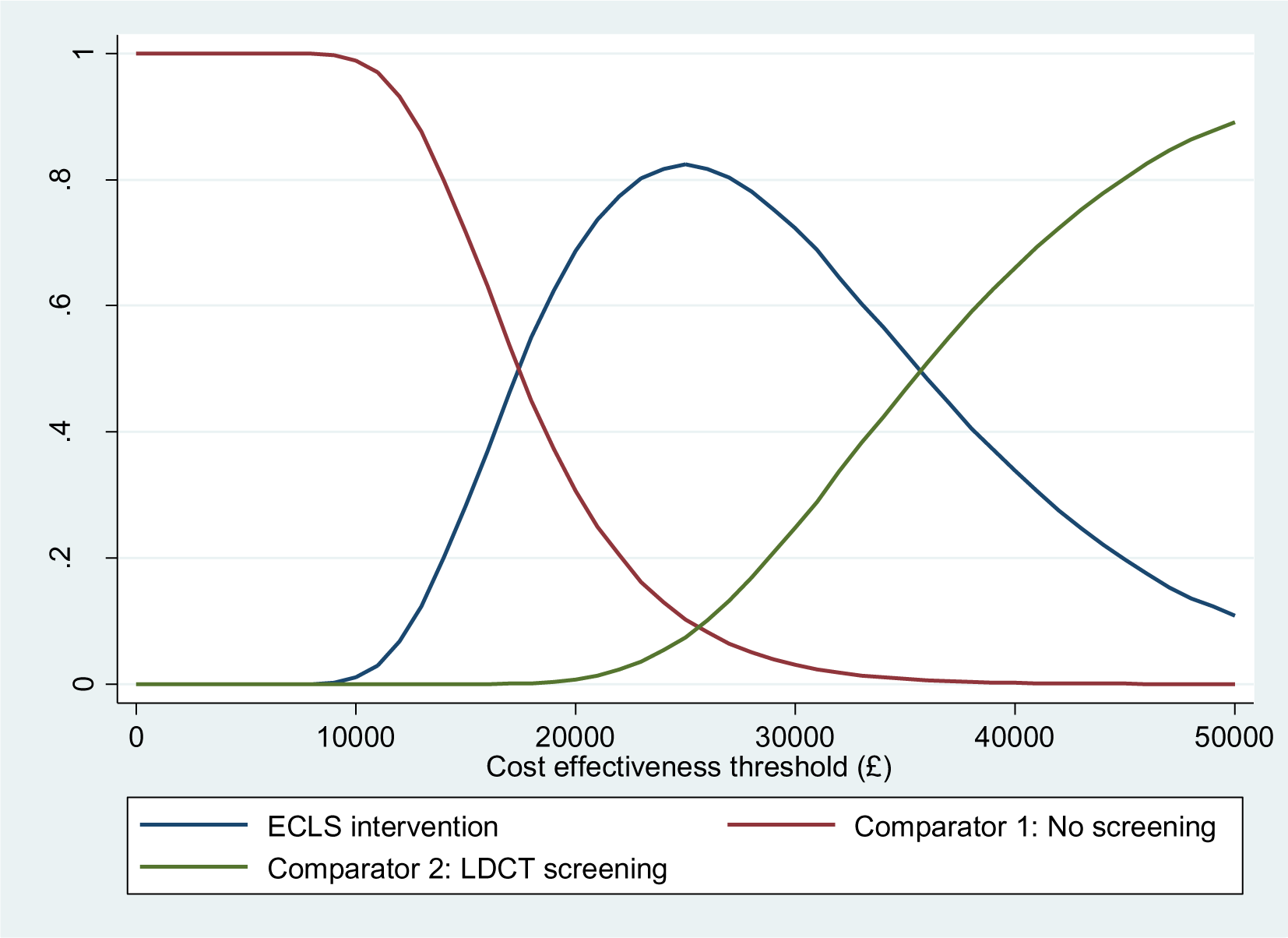
Cost effectiveness acceptability curves

**Fig 3.**
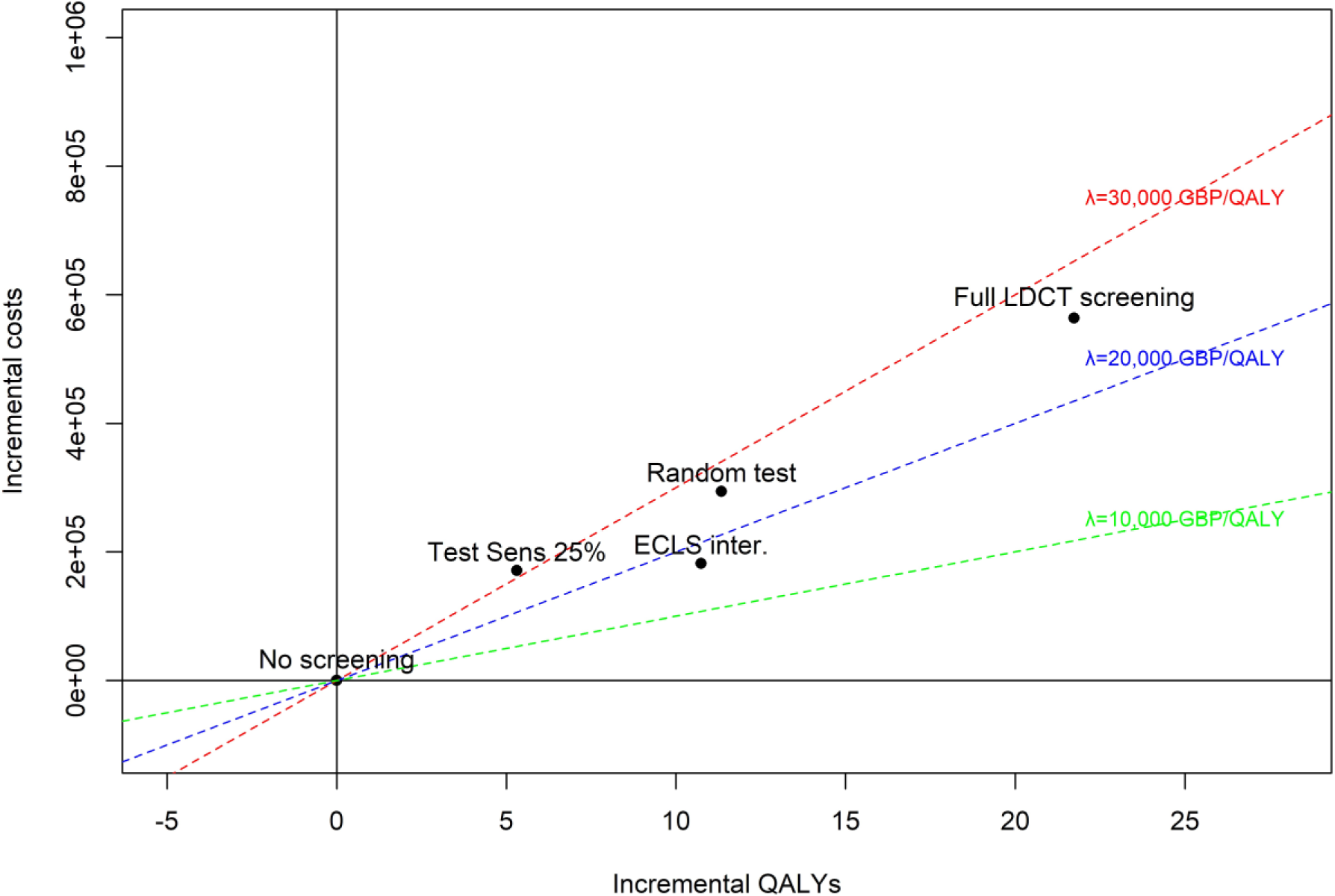
Scenario vs. base case alternatives (ECLS intervention, no screening, and LDCT screening). Scenario 1: sensitivity of early-stage LC set to 25%. Scenario 2: “random test”, sensitivity and 1-specificity set to 52% and cost of test set to zero.

## 4 Discussion

The ECLS trial identified a statistically insignificant higher number of LC cases in the no-screening arm compared to the screening arm, however, a larger proportion of cases were ES in the screening arm compared to the no-screening arm. To address this surprising result, we used modelling techniques to conduct the economic evaluation. The base case analysis estimated a cost per QALY gained of less than £20,000 when comparing the ECLS intervention to a no screening strategy. Only reducing prevalence to 1% or setting the cost of the EarlyCDT®-Lung Test above £190 could make *no screening* the highest NMB alternative at the policy relevant cost-effectiveness thresholds. The base case analysis also concluded that a full LDCT screening programme is not cost-effective unless the prevalence of LC among the target population is assumed 4%. The scenario analysis showed that a screening policy using the EarlyCDT®-Lung Test performs better than a random test given its capacity to differentiate between LC patients and healthy individuals, e.g. 90% of healthy participants saves unnecessary LDCT. These results suggest that the ECLS intervention has the potential to be cost-effective when considering detection of ES LC.

### 4.1 Comparison to other studies

As this is the first health economic evaluation of a diagnostic blood test for LC screening it is not possible to compare results to previous studies, and we were limited to comparing to studies reporting LDCT screening strategies. The National Lung Screening Trial (NLST) in the United States reported a cost of $81,000 per QALY for annual three-year LDCT screening compared to no-screening.[33] The UK Lung-cancer Screening (UKLS) trial reported an estimated ICER of £8,466 (95% CI £5,516 to £12,634) per QALY gained for once-only screening with follow-up if necessary, compared to no screening (follow-up 12-15 months).[7] An analysis based on the methods used in the UKLS trial and evidence from a LC screening pilot in Manchester UK reported an ICER of £10,069. A model also based on the UKLS results demonstrated a lifetime ICER of £28,784 for single screening and £95,292 for triple screening (baseline, 12- and 24-months).[34] These results compare to an average incremental cost per QALY of £25,972 for LDCT screening vs. *no screening* in this study; however this is for 6-monthly LDCT scans compared to once-only screening in the UKLS trial and annual three-year screening in NLST. The cost-effectiveness of LDCT screening has been estimated to change with frequency of screening, age and risk of the target population; in this sense, the estimated cost per QALY gain of an every-year, for ages 55-65, screening program in Spain is closer to the estimates for the 6-monthly screening intervention designed in the ECLS study.[35, 36]

### 4.2 Strengths

The large sample of over 12,000 participants allowed us the best opportunity to find differences in the distribution of LC stages in screening and no-screening arms. The analysis captured the long-term consequences in terms of costs and outcomes of detecting LC at an ES compared to LS. The results highlighted these consequences in terms of lower treatment costs and improved survival and quality of life. The NMB measure allows comparison between alternatives at the policy relevant thresholds. Modelling allows us to explore the scenarios under which the ECLS intervention will be (most) cost-effective by varying the prevalence or the cost of the blood test. The population chosen was slightly younger and had less pack years compared to previous cost-effectiveness analyses of LC screening strategies[33], suggesting that the present study results are a conservative estimate of cost-effectiveness. Specific criticisms of the ECLS study design by Baldwin et al. have been addressed by including no screening and LCDT screening as comparators, as well as including additional alternatives in the scenario analyses[13]. The main criticism was that regular CT scans were offered to test positive participants only; Baldwin et al. suggest that, therefore, generating a test result at random would result in earlier diagnosis in the screening group. To address this criticism, we mimicked random test results using our model; the ECLS study proved to be a more cost-effective alternative than a zero-cost random test. The second criticism was that sensitivity of the EarlyCDT®-Lung Test was overestimated, to address this we reduced the sensitivity of the test to more than half (25% vs. 52% estimated in the ECLS trial); the resulting cost per QALY of this alternative (compared to *no screening*) was just above £30,000. The final criticism was that the EarlyCDT®-Lung Test should be compared to CT scan screening, this was not chosen as a comparator in the ECLS trial as CT scanning was not the standard of care in Scotland when the trial was designed and approved, nor was there capacity to conduct this number of scans. To address this criticism in the economic evaluation we mimicked all participants in the screening arm receiving 6-monthly LDCT scans, again the resulting ICER (vs. no screening) was £25,972 for the base case scenario.

### 4.3 Limitations

The main limitation was lack of resource use data, which led us to use a model-based approach, populating it with a mix of trial data and expert opinion. This may have underestimated the resource use of confirmatory diagnoses as some participants may have had more than one confirmatory diagnostic test, receiving initial negative diagnostic results but needing additional testing after that. Participants may have modified their smoking behaviour due to participation in the study; participants in the no-screening arm may have been more aware of LC symptoms and more likely to seek medical help than had they not been in the study, participants in the test negative group may have felt able to engage in risky behaviour if they felt they were ‘invincible’, and those in the test positive group, without a confirmatory diagnosis, may have changed their smoking behaviour if they felt a positive result was an indication of increased risk of LC diagnosis[37]. However, it is noteworthy that a subgroup analysis of ECLS patients who had nodules detected showed no change in smoking behaviour.[38] We did not include the cost to the NHS of identifying high risk patients and inviting them to screening, the UKLS trial estimated the cost per person of selection and invitation to be £10[7].

The ECLS study is continuing to monitor participants and has five-and ten-year analyses planned.

### 4.4 Conclusion

The base case analysis results estimated that the ECLS intervention is the most cost-effective alternative, with highest probability, when compared to no screening or LDCT screening. This result may change with modifications of the parameters *prevalence of lung cancer* and *EarlyCDT®-Lung Test cost*, suggesting that the three alternatives considered in the main analysis are potentially cost-effective depending on the disease risk of the target population and the cost of testing.

## Statements and Declarations

### Funding

Funding for the ECLS study was received from Oncimmune Ltd and the Scottish Government Health & Social Care Directorate of the Chief Scientist Office (CSO).

### Conflicts of Interest

All authors report grants from Oncimmune Ltd., and grants from Scottish Government Health & Social Care Directorate of the Chief Scientist Office (CSO), during the conduct of the study.

### Availability of data and material

No data is available.

### Ethics approval

The study was conducted in accordance with the principles of Good Clinical Practice and the UK National Research Governance Framework. The University of Dundee and Tayside Health Board co-sponsored the trial, which was registered at ClinicalTrials.gov with identifier NCT01925625. Institutional Review Board approval was provided by the East of Scotland Research Ethics Committee (REC 13/ES/0024).

### Author contributions

FS, FM and AB contributed to study inception and design. Data analysis was undertaken by NM. Model design and analysis was undertaken by JRZ and AB. The first draft of the manuscript was written by JRZ and NM and all authors commented on previous versions of the manuscript. All authors read and approved the final manuscript.

## Data Availability

No data is available.

## Acknowledgements

ECLS trial team, particularly Joana Rocha, Petra Rauchhaus and Pauline Armory.

## Notes

### Clinical Trial

NCT01925625

### Clinical Protocols

https://bmccancer.biomedcentral.com/articles/10.1186/s12885-017-3175-y

