## Supplementary Material for "Health economic evaluation of lung cancer screening using a diagnostic blood test: the Early detection of Cancer of the Lung Scotland (ECLS)"

### Model parameters

Table A1: Description and estimation of model parameters

| Parameter | Description | Estimation and assumptions |
|---|---|---|
| <u>Probabilities</u> | | |
| Prevalence ($p$) | Prevalence of lung cancer (LC). It is the percentage of patients that have LC or will develop LC within the two years from randomisation, i.e. during the ECLS trial period. | Assumed based on ECLS trial protocol[1]. Changed in deterministic sensitivity analysis. |
| $p_{ES}=P(ES\|D+)$ | Relative prevalence of early stage (ES) LC at randomisation point. If all patients with LC were screened during the ECLS trial with a perfect test (100% sensitivity), this would be the percentage of LCs that would be detected at ES. | Calibrated from the sensitivity of the EarlyCDT test and number of true positive cancers detected by the EarlyCDT: <br> $Number\ of\ ES\ cancers = Number\ of\ test\ positive\ ES/Sens\_Test_{ES}$ <br> $Number\ of\ LS\ cancers = Number\ of\ test\ positive\ LS/Sens\_Test_{LS}$. <br> The parameter $P(ES\|D+)$ could be obtained as: <br> $P(ES\|D+) = \frac{Number\ of\ ES\ cancers}{(Number\ of\ ES\ cancers + Number\ of\ LS\ cancers)}$. |

Table A1: Description and estimation of model parameters

| Parameter | Description | Estimation and assumptions |
|---|---|---|
| *$p_{LS}$=P(LS\|D+)=1- $p_{ES}$* | Relative prevalence of late stage (LS) LC at randomisation point. If all patients with LC were screened during the ECLS trial with a perfect test (100% sensitivity), this would be the percentage of LCs that would be detected at LS. | See calibration of *$p_{ES}$ = P(ES\|D+).* |
| Test sensitivity_ES: $P(T^+\|ES)$ | Sensitivity of the EarlyCDT®-Lung test for ES cancer. | Estimated in the ECLS study.[2] |
| Test sensitivity_LS: $P(T^+\|LS)$ | Sensitivity of the EarlyCDT®-Lung test for LS cancer. | Estimated in the ECLS study. [2] |
| *LDCT sensitivity: $P(LDCT^+\|D^+)$* | Sensitivity of the LDCT scanning for any stage cancer. | Assumption. |
| *LDCT specificity: $P(LDCT^-\|D^-)$* | Specificity of the LDCT scanning for any stage cancer. | Assumption. |
| *Opportunistic detection of ES cancer ($Op_{ES}$)* | Probability of oportunistic detection for an ES LC. | Ratio of the number of ES cases detected in the ECLS trial control arm over total number of ES cases expected. |
| *Opportunistic detection of LS cancer ($Op_{LS}$)* | Probability of oportunistic detection for a LS LC. | Ratio of the number of LS cases detected in the ECLS trial control arm over total number of LS cases expected. |
| <u>Costs</u> | | |
| Cost of EarlyCDT-Lung test | Cost of administering the EarlyCDT-Lung, including cost of the test kit and staff cost. | Assumed £106.5 in the base case analysis (15 minutes of nurse time included). Changed in deterministic sensitivity analysis. |
| Diagnostic costs | It comprises the cost incurred for investigations of LC (other than the cost of the EarlyCDT-Lung test). | The next estimations were used for each diagnostic pathway:<br>- TP, FP:<br>o Number of x-ray and LDCT scans as per ECLS trial screening protocol was the average found in the study for participants with a positive test result. |

Table A1: Description and estimation of model parameters

| **Parameter** | **Description** | **Estimation and assumptions** |
|---|---|---|
| | | o Confirmatory diagnostic costs were assumed to be: an x-ray, a contrast LDCT, and the average of either a bronchoscopy or CT guided biopsy.<br>- TP (not LDCT screened)<br>o Confirmatory diagnostic costs as above for TP and FP.<br>- TN (LDCT screened), FN (LDCT screened)<br>o Number of x-ray and LDCT scans according to ECLS trial screening protocol. Estimated as above for TP and FP.<br>- TN, FN: no diagnostic costs assigned.<br><br>See unit costs of diagnostic procedures used in the analysis in Table A2 of this supplementary document. |
| Treatment costs | This is the costs of treatment that will be different for early stage and late stage cancer | Assumed as computed in the UK National Screening Committee (UK-NSC) report authored by the Exeter Test Group and Health Economics Group.[3] See table A7 |
| Proportion of False Negative being treated after screening period | This parameter indicate what percentage of False Negative (i.e. lung cancer non-detected during the two-year ECLS intervention) will be detected before dying. These patients are assumed to be able to receive treatment and incurr in related costs. | Based on *Opportunistic detection of LS cancer* ($Op_{LS}$) and survival function calibrated for LS cancers. See details in this supplementary material "Proportion of False Negative detected after the screening intervention" |
| Proportion of ES LC being treated after recurrence | Proportion of lung cancers, detected at early stage, that recur in a late stage and receive treatment and incur in related costs. | Based on CRUK report "Saving lives, averting costs"[4] |
| <u>Outcomes</u> | | |

Table A1: Description and estimation of model parameters

| Parameter | Description | Estimation and assumptions |
|---|---|---|
| Lead time | This parameter refers to the average time passed between randomisation point and cancer detecction. | The next estimations were used for:<br>- LC detected at ES: average lead time for LC cases with a positive test result in the ECLS trial intervention arm.<br>- LC detected at LS: average lead time for LC cases in the control arm and LC cases with a negative test result in the intervention arm. |
| LE_ES | Life expectancy for ES LC. | The estimation of life expectancy for ES LC has three steps: 1) Estimation of Weibull survival function using digitised K-M curves from the International Association for the Study of LC (IASLC) staging project *[5]*; 2) Calibration of scale parameter $\lambda$ using survival data from the UK; 3) Computation of Life expectancy for lung cancer at early stages by integrating the Weibull function using parameters estimated/calibrated in 1) and 2). See details in this supplementary material "Estimation of life expectancy". |
| LE_LS (non-treated) | Life expectancy for LS LC (non-treated). | The estimation of life expectancy for LS LC (non-treated) has three steps: 1) Estimation of Weibull survival function using digitised K-M curves from the International Association for the Study of LC (IASLC) staging project *[5]*; 2) Calibration of scale parameter $\lambda$ using survival data from the UK; 3) Computation of Life expectancy for lung cancer at late stages by integrating the Weibull function using parameters estimated/calibrated in 1) and 2). See details in this supplementary material "Estimation of life expectancy". |
| LE_LS (treated) | Life expectancy for LS LC (immunotherapy-treated). | The estimation of life expectancy for LS LC (immunotherapy-treated) has four steps: 1) Estimation of Weibull survival function using digitised K-M curves from the International |

Table A1: Description and estimation of model parameters

| Parameter | Description | Estimation and assumptions |
|---|---|---|
| | | Association for the Study of LC (IASLC) staging project *[5]*; 2) Calibration of scale parameter $\lambda$ using survival data from the UK; 3) Adjusting mortality hazard by applying HR estimated in the literature for immunotherapy vs. chemotherapy[6]; 4) Computation of Life expectancy for lung cancer at early and late stages by integrating the Weibull function using parameters estimated/calibrated in 1), 2) and 3). See details in this supplementary material "Estimation of life expectancy". |
| HU_NLC | Health utility for no LC patients | Estimated from ECLS data for participants who did not develop LC. |
| HU_LC_change | Decrement utility for LC in general | Estimated from ECLS data for participants who developed LC. Assumed to be a general utility decrement that applies at the lowest stage of lung cancer. Additional decrements were applied to ES and LS LCs. |
| HU_ES_change | Additional decrement utility for ES LC | Health utilities estimated in Grutters et al. (2010) were used to apply utility decrements for early and late stage of LC. [7] See in this supplementary material "Health utilities". |
| HU_LS_change | Additional decrement utility for LS LC | Health utilities estimated in Grutters et al. (2010) were used to apply utility decrements for early and late stage of LC. [7] See in this supplementary material "Health utilities". |

**Unit costs**

Table A2: Unit costs (2021-22 pounds sterling)

| Resource use | Unit cost (£) | Source | Details |
|---|---|---|---|
| Blood test kit | 95.00 | Oncimmune | $124 per kit |
| Test administration | 11.5 | PSSRU 2021/22 | 15 minutes of nurse time (GP surgery) (£46 per hour) |
| LDCT scan one area | 95.5 | NHS reference costs 2017/18 and updated to 2022 using NHS Cost Inflation Index for prices | CT of one area, no contrast RD20A in diagnostic imaging tab, outpatient |
| LDCT scan two areas | 113.5 | NHS reference costs 2017/18 and updated to 2022 using NHS Cost Inflation Index for prices | CT scan of two areas with contrast (chest and abdomen) RD24Z, diagnostic imaging tab, outpatient |
| X-ray | 64.5 | ISD costs 2017/18 and updated to 2022 using NHS Cost Inflation Index for prices | Other radiology R120X |
| Bronchoscopy | 954.73 | NHS reference costs 2021/22 | DZ69A respiratory medicine service diag bronch OP procedures |
| CT guided biopsy | 1547.73 | NHS reference costs 2021/22 | DZ63C daycase major thoracic procedure 19+ CC score 0-2 |

**Estimation of life expectancy at diagnosis**

A Weibull model was used to compute life expectancy from diagnosis for each LC at stage $i$:

$$\int_0^\infty S_i(t) = \int_0^\infty \exp\left(-\lambda_i t^p\right). \tag{1}$$

Where $S_i(t)$ is the survival function and t=0 is the moment of diagnosis. Parameters $p$ and $\lambda_i$ were first estimated using digitized data from K-M curves published by the IASLC staging project.*[5]* Then, parameter $\lambda_i$ was calibrated for each stage I, II, III and IV using five-year survival figures from England data:

$$\lambda_i = \frac{-\log\left(S_i(5)\right)}{5^p}. \tag{2}$$

Where log is the natural logarithm and $S_i(5)$ is the five-year survival figure using data available from England.

The estimation of life expectancy has three steps: 1) Estimation of Weibull survival function using digitised K-M curves from the International Association for the Study of LC (IASLC) staging project*[5]*; 2) Calibration of scale parameter $\lambda$ using survival data from the UK; 3) Computation of Life expectancy for each stage by integrating the Weibull function using parameters estimated/calibrated in 1) and 2).

The estimation of life expectancy for LS LC (immunotherapy-treated) is computed seemingly but applying a HR, estimated in the literature for immunotherapy vs. chemotherapy, to the scale parameters $\lambda$. [6]

<u>Estimation of Weibull survival function</u>

The IASLC staging project presented data for 17,509 observations representing patients in different LC stages (see figure 2A in *[5]*). Data covers a period of 72 months from diagnosis. Given that no information is shown on censoring during this period, all censoring was assumed to happen at the end of the period.

Table A3. Weibull survival function estimation

| | Coef. | S.E. |
|---|---|---|
| Log($\lambda$): | | |
| Stage IB | 0.617 | 0.041 |
| Stage IIA | 1.040 | 0.050 |
| Stage IIB | 1.198 | 0.053 |
| Stage IIIA | 1.520 | 0.033 |
| Stage IIIB | 2.065 | 0.034 |
| Stage IV | 2.654 | 0.044 |
| Cons (Stage IA) | -2.875 | 0.029 |
| Log($p$) | -0.143 | 0.009 |
| *N* | 17,509 | |

Note: digitized data from the IASLC staging project used

Calibration of scale parameter λ to UK data

Data from England NHS-Digital publication "*Cancer Survival in England, cancers diagnosed 2015 to 2019, followed up to 2020*" is shown in Table A4.[8]

Table A4. 5-year age-standardised survival ($S_i(5)$) for LC. England NHS-Digital

| Stage 1 | Stage 2 | Stage 3 | Stage 4 |
|---|---|---|---|
| 0.609 | 0.387 | 0.147 | 0.039 |

The average of the scale parameter $\lambda_i$ for each stage $i$ is calibrated to meet five-year survival figures. A standard error for the scale parameter is applied for each scale parameter using estimations in Table A3.

Table A5. Calibration of Weibull scale parameter ($\lambda$)

| | Log($\lambda$) | | | |
|---|---|---|---|---|
| | Stage 1 | Stage 2 | Stage 3 | Stage 4 |
| Average | -1.959 | -1.322 | -0.667 | -0.131 |
| S.E. | 0.029 | 0.045 | 0.024 | 0.035 |

Note. Standard errors taken from estimations of $\lambda$ for each stage in Table A3.

Also, the HR used to adjust the mortality hazard for immunotherapy-treated LS LC is 0.62 (95% CI, 0.48 to 0.81).[6] A lognormal distribution, $logHR \sim N(-0.478, 0.135)$, has been used to account for uncertainty in the HR, where the standard deviation 0.135 has been calibrated to be consistent with the 95% confidence interval in the HR.

Computation of Life expectancy

Life expectancy is computed by integrating the Weibull survival function using the estimated shape parameter and calibrated scale parameter (adjusted for immunotherapy treatment as appropriate). The results are shown in Table A6 below. The average of life expectancy for stage 1 and stage 2 are applied to cancers detected at early stage in our model. Late-stage cancers are assigned the average for stage 3 and stage 4.

Table A6. Life expectancy for each LC stage

| | Stage 1 | Stage 2 | Stage 3 (non-treated) | Stage 4 (non-treated) | Stage 3 (immun-treated) | Stage 4 (immun-treated) |
|---|---|---|---|---|---|---|
| Average | 12.047 | 5.703 | 2.538 | 1.384 | 4.405 | 2.401 |
| S.E. | 0.430 | 0.271 | 0.0758 | 0.0571 | 0.697 | 0.396 |

**Treatment costs**

The CRUK report "Saving lives, averting costs" presented treatment costs for LC patients according to their disease stage. [4] Early-stage LC treatment costs were computed as the average of stage I and II.

Table A7: Treatment costs (£) at diagnosis from the UK National Screening Committee (UK-NSC) report

| Type of treatment | Lung cancer stage | | | |
|---|---|---|---|---|
| | I | II | III | IV |
| Surgery | 2982 | 2895 | 847 | 549 |
| Radio Therapy (RT) | 909 | 791 | 472 | 730 |
| Chemo-rad | | | 1817 | |
| Chemo Therapy (CT) | | | | 782 |
| CT doublet | | 587 | 105 | 393 |
| Immunotherapy | | | 8426 | 2640 |
| Chemo-immunotherapy | | | 3585 | 6692 |
| TKI | | | 1334 | 3772 |
| Follow-up | 271 | 217 | 162 | 108 |
| Total | 4162 | 4490 | 16748 | 15666 |

Late-stage lung cancers were assigned the average of treatment cost for stage III and IV. Early-stage lung cancers were assigned the average of treatment costs for stage I and II.

**Proportion of False Negative detected after the screening intervention.**

In the model, False Negative patients did not receive a correct diagnosis during the period of the ECLS intervention and therefore were assumed to have been detected later at an advanced stage. But not all of them will be diagnosed before dying. It was assumed that they would be detected according to opportunistic detection rate for LS LC. Using the opportunistic detection rate during the two years of the ECLS intervention (0.7208), we can compute the one-year probability of detection as $opp = 1 - (1 - 0.7208)^{1/2}$. The proportion of False Negative patients detected before dying can be computed as:

$$\sum_{t=2}^{t=T} FN_{\mathrm{det}(t)} = \sum_{t=2}^{t=T}\left(opp \times \left[S(t)\left(1 - \sum_{i=2}^{t} FN_{\mathrm{det}(i)}\right)\right]\right),$$

where $t$ is the index for year since end of screening intervention at year 2, T is the year where all the False Negative are dead, $FN_{\mathrm{det}(t)}$ are the False Negative patients detected in year t, and $S(t)$ is the proportion of False Negative that survives to year t. Using the Weibull survival function for LS LC we obtain $\sum_{t=2}^{t=T} FN_{\text{det}(t)} = 0.3885$. The average year of detection, when treatment is expected to happen for these patients, is t=3.

**Health utilities**

Health utilities estimated in Grutters et al. (2010) were used to apply utility decrements different stages of LC. [7] Based on these figures additional utility decrements in the model were 0.03 and 0.07 for early and late stage LC.

Table A8. Utility decrements based on Grutters et al. (2010)

| | Original table | | | Model parameters | |
|---|---|---|---|---|---|
| Stage | N | Mean | Sd | Utility decrements w.r.t. stage I | S.E. |
| I | 105 | 0.77 | 0.26 | 0 | 0.025 |
| II | 39 | 0.74 | 0.22 | -0.03 | 0.035 |
| III | 99 | 0.7 | 0.29 | -0.07 | 0.029 |
| IV | 2 | 0.86 | 0.19 | -0.07 | 0.029 |

Note: Given lack of data, model parameters for stage IV are assumed the same as for stage III.

**Cohort counts for each model pathway (baseline results). Cohort of 1,000 patients**

Table A9. Cohort counts for each model pathway. Baseline results. 1,000 patients

| | Endpoint (diagnosis pathway and cancer stage at diagnosis) | | | | | | | | | | |
|---|---|---|---|---|---|---|---|---|---|---|---|
| Strategy | FP (NLC) | TN LDCT-screened (NLC) | TN (NLC) | TP (ES) | FN LDCT-screened (LS) | TP NotLDCT-screened (ES) | FN (LS) | TP (LS) | FN LDCT-screened (LS) | TP NotLDCT-screened (LS) | FN (LS) |
| ECLS intervention | 0 | 94.2 | 886 | 4.29 | 0 | 1.48 | 2.44 | 2.14 | 0 | 6.95 | 2.69 |
| Comparator 1: No screening | 0 | 0 | 980 | 0 | 0 | 3.1 | 5.11 | 0 | 0 | 8.5 | 3.29 |
| Comparator 2: LDCT screening | 0 | 980 | 0 | 8.21 | 0 | 0 | 0 | 11.8 | 0 | 0 | 0 |

TN – true negative; TP – true positive; FN – false negative; FP – false positive; LDCT – low-dose computed tomography; LC – lung cancer; NLC – no lung cancer; ES – early stage; LS – late-stage

**Deterministic sensitivity analysis**

| Parameter assumption | ECLS intervention | Comp. 1: No screening | Comp. 2: LDCT screening |
|---|---|---|---|
| *EarlyCDT cost (£201.5)* | | | |
| Costs (£) | 509,102 | 232,421 | 796,016 |
| QALYs | 8,570.4 | 8,559.7 | 8,581.4 |
| NMB (λ=£20,000) | 170,899,000 | 170,961,000 | 170,832,000 |
| NMB (λ=£30,000) | 256,603,000 | 256,557,000 | 256,645,000 |
| ΔCosts (£) | | 276,681 | -286,915 |
| ΔQALYs | | 10.7 | -11.0 |
| ΔNMB (λ=£20,000) | | -61,821 | 67,095 |
| ΔNMB (λ=£30,000) | | 45,609 | -42,816 |
| *EarlyCDT cost (£59)* | | | |
| Costs (£) | 366,602 | 232,421 | 796,016 |
| QALYs | 8,570.4 | 8,559.7 | 8,581.4 |
| NMB (λ=£20,000) | 171,041,000 | 170,961,000 | 170,832,000 |
| NMB (λ=£30,000) | 256,745,000 | 256,557,000 | 256,645,000 |
| ΔCosts (£) | | 134,181 | -429,415 |
| ΔQALYs | | 10.7 | -11.0 |
| ΔNMB (λ=£20,000) | | 80,679 | 209,595 |
| ΔNMB (λ=£30,000) | | 188,109 | 99,685 |
| *Lead time (0.4443)* | | | |
| Costs (£) | 414,102 | 232,421 | 796,016 |
| QALYs | 8,565.2 | 8,553.4 | 8,577.1 |
| NMB (λ=£20,000) | 170,889,000 | 170,836,000 | 170,746,000 |
| NMB (λ=£30,000) | 256,541,000 | 256,370,000 | 256,517,000 |
| ΔCosts (£) | | 181,681 | -381,915 |
| ΔQALYs | | 11.7 | -11.9 |
| ΔNMB (λ=£20,000) | | 53,219 | 143,294 |
| ΔNMB (λ=£30,000) | | 170,669 | 23,985 |
| *Lead time (2)* | | | |
| Costs (£) | 414,102 | 232,421 | 796,016 |
| QALYs | 8,578.4 | 8,569.2 | 8,588.0 |
| NMB (λ=£20,000) | 171,154,000 | 171,152,000 | 170,963,000 |
| NMB (λ=£30,000) | 256,938,000 | 256,844,000 | 256,842,000 |

| Parameter assumption | ECLS intervention | Comp. 1: No screening | Comp. 2: LDCT screening |
|---|---|---|---|
| ΔCosts (£) | | 181,681 | -381,915 |
| ΔQALYs | | 9.2 | -9.5 |
| ΔNMB (λ=£20,000) | | 2,359 | 191,074 |
| ΔNMB (λ=£30,000) | | 94,379 | 95,655 |
| *Opportunistic detection of ES cancer (0)* | | | |
| Costs (£) | 403,428 | 210,103 | 796,016 |
| QALYs | 8,564.6 | 8,547.5 | 8,581.4 |
| NMB (λ=£20,000) | 170,888,000 | 170,740,000 | 170,832,000 |
| NMB (λ=£30,000) | 256,534,000 | 256,215,000 | 256,645,000 |
| ΔCosts (£) | | 193,325 | -392,588 |
| ΔQALYs | | 17.1 | -16.8 |
| ΔNMB (λ=£20,000) | | 148,175 | 56,688 |
| ΔNMB (λ=£30,000) | | 318,925 | -111,262 |
| *Opportunistic detection of ES cancer (1)* | | | |
| Costs (£) | 431,673 | 269,162 | 796,016 |
| QALYs | 8,580.0 | 8,579.6 | 8,581.4 |
| NMB (λ=£20,000) | 171,167,000 | 171,323,000 | 170,832,000 |
| NMB (λ=£30,000) | 256,967,000 | 257,120,000 | 256,645,000 |
| ΔCosts (£) | | 162,512 | -364,343 |
| ΔQALYs | | 0.3 | -1.4 |
| ΔNMB (λ=£20,000) | | -156,132 | 335,623 |
| ΔNMB (λ=£30,000) | | -152,942 | 321,263 |
| *Opportunistic detection of LS cancer (0)* | | | |
| Costs (£) | 335,141 | 135,913 | 796,016 |
| QALYs | 8,566.7 | 8,555.1 | 8,581.4 |
| NMB (λ=£20,000) | 170,999,000 | 170,966,000 | 170,832,000 |
| NMB (λ=£30,000) | 256,665,000 | 256,518,000 | 256,645,000 |
| ΔCosts (£) | | 199,228 | -460,875 |
| ΔQALYs | | 11.6 | -14.7 |
| ΔNMB (λ=£20,000) | | 32,112 | 166,895 |

| Parameter assumption | ECLS intervention | Comp. 1: No screening | Comp. 2: LDCT screening |
|---|---|---|---|
| ΔNMB (λ=£30,000) | | 147,782 | 19,905 |
| *Opportunistic detection of LS cancer (1)* | | | |
| Costs (£) | 444,684 | 269,799 | 796,016 |
| QALYs | 8,571.8 | 8,561.4 | 8,581.4 |
| NMB (λ=£20,000) | 170,992,000 | 170,958,000 | 170,832,000 |
| NMB (λ=£30,000) | 256,710,000 | 256,572,000 | 256,645,000 |
| ΔCosts (£) | | 174,885 | -351,332 |
| ΔQALYs | | 10.4 | -9.6 |
| ΔNMB (λ=£20,000) | | 33,595 | 160,232 |
| ΔNMB (λ=£30,000) | | 137,835 | 64,682 |
| *Prob. of treatment undetected LC (*0.19*)* | | | |
| Costs (£) | 399,194 | 208,039 | 796,016 |
| QALYs | 8,569.5 | 8,558.2 | 8,581.4 |
| NMB (λ=£20,000) | 170,991,000 | 170,955,000 | 170,832,000 |
| NMB (λ=£30,000) | 256,685,000 | 256,537,000 | 256,645,000 |
| ΔCosts (£) | | 191,156 | -396,822 |
| ΔQALYs | | 11.3 | -11.9 |
| ΔNMB (λ=£20,000) | | 35,204 | 158,882 |
| ΔNMB (λ=£30,000) | | 148,384 | 39,912 |
| *Prob. of treatment undetected LC (*0.76*)* | | | |
| Costs (£) | 441,990 | 278,034 | 796,016 |
| QALYs | 8,572.1 | 8,562.4 | 8,581.4 |
| NMB (λ=£20,000) | 170,999,000 | 170,970,000 | 170,832,000 |
| NMB (λ=£30,000) | 256,720,000 | 256,594,000 | 256,645,000 |
| ΔCosts (£) | | 163,956 | -354,027 |
| ΔQALYs | | 9.7 | -9.3 |
| ΔNMB (λ=£20,000) | | 29,504 | 167,847 |
| ΔNMB (λ=£30,000) | | 126,234 | 74,757 |
| *Discount rate (6%)* | | | |
| Costs (£) | 405,102 | 223,735 | 780,617 |
| QALYs | 7,615.0 | 7,605.7 | 7,624.6 |

| Parameter assumption | ECLS intervention | Comp. 1: No screening | Comp. 2: LDCT screening |
|---|---|---|---|
| NMB (λ=£20,000) | 151,895,000 | 151,890,000 | 151,712,000 |
| NMB (λ=£30,000) | 228,045,000 | 227,946,000 | 227,958,000 |
| ΔCosts (£) | | 181,367 | -375,515 |
| ΔQALYs | | 9.3 | -9.6 |
| ΔNMB (λ=£20,000) | | 5,393 | 183,275 |
| ΔNMB (λ=£30,000) | | 98,773 | 87,155 |
| *Discount rate (0%)* | | | |
| Costs (£) | 427,898 | 245,745 | 819,239 |
| QALYs | 10,284.4 | 10,271.2 | 10,297.8 |
| NMB (λ=£20,000) | 205,261,000 | 205,179,000 | 205,137,000 |
| NMB (λ=£30,000) | 308,105,000 | 307,891,000 | 308,115,000 |
| ΔCosts (£) | | 182,153 | -391,341 |
| ΔQALYs | | 13.2 | -13.4 |
| ΔNMB (λ=£20,000) | | 81,847 | 123,341 |
| ΔNMB (λ=£30,000) | | 213,847 | -10,659 |
| *Prevalence (1%)* | | | |
| Costs (£) | 283,707 | 116,210 | 641,390 |
| QALYs | 8,620.1 | 8,614.7 | 8,625.6 |
| NMB (λ=£20,000) | 172,118,000 | 172,178,000 | 171,870,000 |
| NMB (λ=£30,000) | 258,319,000 | 258,325,000 | 258,126,000 |
| ΔCosts (£) | | 167,497 | -357,683 |
| ΔQALYs | | 5.4 | -5.5 |
| ΔNMB (λ=£20,000) | | -60,077 | 247,763 |
| ΔNMB (λ=£30,000) | | -6,367 | 192,803 |
| *Prevalence (4%)* | | | |
| Costs (£) | 674,891 | 464,841 | 1,105,270 |
| QALYs | 8,471.0 | 8,449.5 | 8,493.0 |
| NMB (λ=£20,000) | 168,745,000 | 168,526,000 | 168,755,000 |
| NMB (λ=£30,000) | 253,456,000 | 253,021,000 | 253,685,000 |
| ΔCosts (£) | | 210,050 | -430,378 |
| ΔQALYs | | 21.5 | -22.0 |
| ΔNMB (λ=£20,000) | | 219,670 | -9,282 |

| Parameter assumption | ECLS intervention | Comp. 1: No screening | Comp. 2: LDCT screening |
|---|---|---|---|
| ΔNMB (λ=£30,000) | | 434,530 | -229,112 |
| *Relative prevalence ES (0.25)* | | | |
| Costs (£) | 425,855 | 250,008 | 810,712 |
| QALYs | 8,562.9 | 8,556.1 | 8,570.5 |
| NMB (λ=£20,000) | 170,832,000 | 170,873,000 | 170,600,000 |
| NMB (λ=£30,000) | 256,461,000 | 256,434,000 | 256,305,000 |
| ΔCosts (£) | | 175,847 | -384,857 |
| ΔQALYs | | 6.8 | -7.6 |
| ΔNMB (λ=£20,000) | | -40,827 | 231,977 |
| ΔNMB (λ=£30,000) | | 26,683 | 155,537 |
| *Relative prevalence ES (0.75)* | | | |
| Costs (£) | 389,289 | 195,292 | 764,991 |
| QALYs | 8,586.2 | 8,567.1 | 8,604.3 |
| NMB (λ=£20,000) | 171,335,000 | 171,146,000 | 171,321,000 |
| NMB (λ=£30,000) | 257,198,000 | 256,817,000 | 257,364,000 |
| ΔCosts (£) | | 193,997 | -375,702 |
| ΔQALYs | | 19.2 | -18.1 |
| ΔNMB (λ=£20,000) | | 189,403 | 14,542 |
| ΔNMB (λ=£30,000) | | 381,103 | -166,038 |
| *Prob. recurrence (*0.25*)* | | | |
| Costs (£) | 393,014 | 221,077 | 765,997 |
| QALYs | 8,569.3 | 8,559.1 | 8,579.8 |
| NMB (λ=£20,000) | 170,993,000 | 170,960,000 | 170,830,000 |
| NMB (λ=£30,000) | 256,686,000 | 256,551,000 | 256,629,000 |
| ΔCosts (£) | | 171,938 | -372,983 |
| ΔQALYs | | 10.2 | -10.5 |
| ΔNMB (λ=£20,000) | | 32,762 | 162,463 |
| ΔNMB (λ=£30,000) | | 135,112 | 57,203 |
| *Prob. recurrence (*0.75*)* | | | |
| Costs (£) | 435,189 | 243,764 | 826,035 |
| QALYs | 8,571.5 | 8,560.2 | 8,582.9 |
| NMB (λ=£20,000) | 170,994,000 | 170,961,000 | 170,832,000 |

| Parameter assumption | ECLS intervention | Comp. 1: No screening | Comp. 2: LDCT screening |
|---|---|---|---|
| NMB (λ=£30,000) | 256,709,000 | 256,563,000 | 256,661,000 |
| ΔCosts (£) | | 191,425 | -390,846 |
| ΔQALYs | | 11.2 | -11.4 |
| ΔNMB (λ=£20,000) | | 33,295 | 161,986 |
| ΔNMB (λ=£30,000) | | 145,655 | 47,556 |
| *Test sensitivity_ES* (0.25*)* | | | |
| Costs (£) | 403,257 | 232,421 | 796,016 |
| QALYs | 8,565.0 | 8,559.7 | 8,581.4 |
| NMB (λ=£20,000) | 170,896,000 | 170,961,000 | 170,832,000 |
| NMB (λ=£30,000) | 256,546,000 | 256,557,000 | 256,645,000 |
| ΔCosts (£) | | 170,836 | -392,759 |
| ΔQALYs | | 5.3 | -16.4 |
| ΔNMB (λ=£20,000) | | -64,556 | 64,359 |
| ΔNMB (λ=£30,000) | | -11,416 | -99,841 |
| *Test sensitivity_ES* (0.75*)* | | | |
| Costs (£) | 423,211 | 232,421 | 796,016 |
| QALYs | 8,575.0 | 8,559.7 | 8,581.4 |
| NMB (λ=£20,000) | 171,076,000 | 170,961,000 | 170,832,000 |
| NMB (λ=£30,000) | 256,825,000 | 256,557,000 | 256,645,000 |
| ΔCosts (£) | | 190,791 | -372,805 |
| ΔQALYs | | 15.3 | -6.4 |
| ΔNMB (λ=£20,000) | | 115,269 | 244,185 |
| ΔNMB (λ=£30,000) | | 268,299 | 179,875 |
| *Test sensitivity_LS* (0.09*)* | | | |
| Costs (£) | 409,817 | 232,421 | 796,016 |
| QALYs | 8,570.2 | 8,559.7 | 8,581.4 |
| NMB (λ=£20,000) | 170,995,000 | 170,961,000 | 170,832,000 |
| NMB (λ=£30,000) | 256,697,000 | 256,557,000 | 256,645,000 |
| ΔCosts (£) | | 177,397 | -386,199 |
| ΔQALYs | | 10.6 | -11.2 |
| ΔNMB (λ=£20,000) | | 34,243 | 163,159 |
| ΔNMB (λ=£30,000) | | 140,063 | 51,639 |

| Parameter assumption | ECLS intervention | Comp. 1: No screening | Comp. 2: LDCT screening |
|---|---|---|---|
| *Test sensitivity_LS* (0.36) | | | |
| Costs (£) | 422,415 | 232,421 | 796,016 |
| QALYs | 8,570.7 | 8,559.7 | 8,581.4 |
| NMB (λ=£20,000) | 170,992,000 | 170,961,000 | 170,832,000 |
| NMB (λ=£30,000) | 256,699,000 | 256,557,000 | 256,645,000 |
| ΔCosts (£) | | 189,995 | -373,601 |
| ΔQALYs | | 11.1 | -10.7 |
| ΔNMB (λ=£20,000) | | 31,125 | 160,041 |
| ΔNMB (λ=£30,000) | | 141,685 | 53,261 |
| *Test specificity* (0.5) | | | |
| Costs (£) | 606,740 | 232,421 | 796,016 |
| QALYs | 8,570.4 | 8,559.7 | 8,581.4 |
| NMB (λ=£20,000) | 170,801,000 | 170,961,000 | 170,832,000 |
| NMB (λ=£30,000) | 256,505,000 | 256,557,000 | 256,645,000 |
| ΔCosts (£) | | 374,319 | -189,276 |
| ΔQALYs | | 10.7 | -11.0 |
| ΔNMB (λ=£20,000) | | -159,459 | -30,544 |
| ΔNMB (λ=£30,000) | | -52,029 | -140,454 |
| *Test specificity (1)* | | | |
| Costs (£) | 368,226 | 232,421 | 796,016 |
| QALYs | 8,570.4 | 8,559.7 | 8,581.4 |
| NMB (λ=£20,000) | 171,040,000 | 170,961,000 | 170,832,000 |
| NMB (λ=£30,000) | 256,744,000 | 256,557,000 | 256,645,000 |
| ΔCosts (£) | | 135,805 | -427,790 |
| ΔQALYs | | 10.7 | -11.0 |
| ΔNMB (λ=£20,000) | | 79,055 | 207,970 |
| ΔNMB (λ=£30,000) | | 186,485 | 98,060 |
| *Health utility for no LC* (0.5) | | | |
| Costs (£) | 414,102 | 232,421 | 796,016 |
| QALYs | 4,951.9 | 4,946.4 | 4,957.5 |
| NMB (λ=£20,000) | 98,623,300 | 98,694,900 | 98,352,900 |
| NMB (λ=£30,000) | 148,142,000 | 148,159,000 | 147,927,000 |

| Parameter assumption | ECLS intervention | Comp. 1: No screening | Comp. 2: LDCT screening |
|---|---|---|---|
| ΔCosts (£) | | 181,681 | -381,915 |
| ΔQALYs | | 5.5 | -5.6 |
| ΔNMB (λ=£20,000) | | -71,621 | 270,355 |
| ΔNMB (λ=£30,000) | | -16,591 | 214,575 |
| *Health utility for no LC (1)* | | | |
| Costs (£) | 414,102 | 232,421 | 796,016 |
| QALYs | 9,920.9 | 9,908.2 | 9,933.9 |
| NMB (λ=£20,000) | 198,004,000 | 197,932,000 | 197,882,000 |
| NMB (λ=£30,000) | 297,213,000 | 297,014,000 | 297,222,000 |
| ΔCosts (£) | | 181,681 | -381,915 |
| ΔQALYs | | 12.7 | -13.0 |
| ΔNMB (λ=£20,000) | | 72,299 | 121,695 |
| ΔNMB (λ=£30,000) | | 199,289 | -8,416 |
| *Health utility for no LC (1)* | | | |
| Costs (£) | 409,817 | 232,421 | 796,016 |
| QALYs | 8,570.2 | 8,559.7 | 8,581.4 |
| NMB (λ=£20,000) | 170,995,000 | 170,961,000 | 170,832,000 |
| NMB (λ=£30,000) | 256,697,000 | 256,557,000 | 256,645,000 |
| ΔCosts (£) | | 177,397 | -386,199 |
| ΔQALYs | | 10.6 | -11.2 |
| ΔNMB (λ=£20,000) | | 34,243 | 163,159 |
| ΔNMB (λ=£30,000) | | 140,063 | 51,639 |
| *Health utility for no LC (1)* | | | |
| Costs (£) | 422,415 | 232,421 | 796,016 |
| QALYs | 8,570.7 | 8,559.7 | 8,581.4 |
| NMB (λ=£20,000) | 170,992,000 | 170,961,000 | 170,832,000 |
| NMB (λ=£30,000) | 256,699,000 | 256,557,000 | 256,645,000 |
| ΔCosts (£) | | 189,995 | -373,601 |
| ΔQALYs | | 11.1 | -10.7 |
| ΔNMB (λ=£20,000) | | 31,125 | 160,041 |
| ΔNMB (λ=£30,000) | | 141,685 | 53,261 |
| *Health utility for no LC (1)* | | | |
| Costs (£) | 440,808 | 232,421 | 1,073,720 |

| Parameter assumption | ECLS intervention | Comp. 1: No screening | Comp. 2: LDCT screening |
|---|---|---|---|
| QALYs | 8,570.4 | 8,559.7 | 8,581.4 |
| NMB (λ=£20,000) | 170,967,000 | 170,961,000 | 170,554,000 |
| NMB (λ=£30,000) | 256,671,000 | 256,557,000 | 256,368,000 |
| ΔCosts (£) | | 208,387 | -632,907 |
| ΔQALYs | | 10.7 | -11.0 |
| ΔNMB (λ=£20,000) | | 6,473 | 413,087 |
| ΔNMB (λ=£30,000) | | 113,903 | 303,177 |
| *Health utility for no LC (1)* | | | |
| Costs (£) | 424,784 | 232,421 | 907,096 |
| QALYs | 8,570.4 | 8,559.7 | 8,581.4 |
| NMB (λ=£20,000) | 170,983,000 | 170,961,000 | 170,721,000 |
| NMB (λ=£30,000) | 256,687,000 | 256,557,000 | 256,534,000 |
| ΔCosts (£) | | 192,364 | -482,312 |
| ΔQALYs | | 10.7 | -11.0 |
| ΔNMB (λ=£20,000) | | 22,497 | 262,492 |
| ΔNMB (λ=£30,000) | | 129,927 | 152,582 |
